# Sensitive periods for exposure to indoor air pollutants and psychosocial factors in association with symptoms of psychopathology at school-age in a South African birth cohort

**DOI:** 10.1101/2023.08.08.23293825

**Authors:** Grace M. Christensen, Michele Marcus, Aneesa Vanker, Stephanie M. Eick, Susan Malcolm-Smith, Andrew D.A.C. Smith, Erin C. Dunn, Shakira F. Suglia, Howard H. Chang, Heather J. Zar, Dan J. Stein, Anke Hüls

## Abstract

**Background:** Gestation and the first few months of life are important periods for brain development. During these periods, exposure to environmental toxicants and psychosocial stressors are particularly harmful and may impact brain development. Specifically, exposure to indoor air pollutants (IAP) and psychosocial factors (PF) during these sensitive periods has been shown to predict childhood psychopathology.

**Objectives:** This study aims to investigate sensitive periods for the individual and joint effects of IAP and PF on childhood psychopathology at 6.5 years.

**Methods:** We analyzed data from the Drakenstein Child Health Study (N=599), a South African birth cohort. Exposure to IAP and PF was measured during the second trimester of pregnancy and 4 months postpartum. The outcome of childhood psychopathology was assessed at 6.5 years old using the Childhood Behavior Checklist (CBCL). We investigated individual effects of either pre-or postnatal exposure to IAP and PF on CBCL scores using adjusted linear regression models, and joint effects of these exposures using quantile g-computation and self-organizing maps (SOM). To identify possible sensitive periods, we used a structured life course modeling approach (SLCMA) as well as exposure mixture methods (quantile g-computation and SOM).

**Results:** Prenatal exposure to IAP or PFs, as well as the total prenatal mixture assessed using quantile g-computation, were associated with increased psychopathology. SLCMA and SOM models also indicated that the prenatal period is a sensitive period for IAP exposure on childhood psychopathology. Depression and alcohol were associated in both the pre-and postnatal period, while CO was associated with the postnatal period.

**Discussion:** Pregnancy may be a sensitive period for the effect of indoor air pollution on childhood psychopathology. Exposure to maternal depression and alcohol in both periods was also associated with psychopathology. Determining sensitive periods of exposure is vital to ensure effective interventions to reduce childhood psychopathology.

## Introduction

Childhood psychopathology, including anxiety, attention-deficit/hyperactivity disorder (ADHD), oppositional defiant disorder, and depression, can cause problems for children in all aspects of life^1^. Previous studies have found that children who experience elevated symptoms of psychopathology are at a higher risk of developing mental health problems as adults^2^. The two major categories of psychopathology symptoms and disorders are internalizing and externalizing. Externalizing behaviors, such as aggression, delinquency, and hyperactivity are displayed outwardly and reflect the behavior towards the environment^3^. In contrast, internalizing behaviors, such as depression, anxiety, somatic complaints, and suicidal ideation, are directed inward and reflect the child’s emotional and psychological state^4^.There is a gap in knowledge on how to prevent psychopathology in children^2^. Bridging this gap through identification of modifiable risk factors has potential to reduce the burden of mental health problems in childhood.

Air pollution may be one modifiable risk factor, shown in both animal experiments and human observational studies to affect the central nervous system (CNS) and elements of behavior and psychopathology^5–9^. Additionally, exposure to psychosocial factors and their determinants which may induce stress, such as socioeconomic status, substance use, violence, and psychological distress, also affects child behavioral development and psychopathology. Animal studies have shown prenatal stress affects behavior in rats ^10–12^, and there is growing epidemiological evidence for pre-and postnatal psychosocial stress’ effects on child cognitive development^13^. However, few studies have examined the combined association of air pollutants and psychosocial factors on psychopathology.

While exposure to both indoor air pollution and psychosocial factors have been associated with childhood psychopathology separately^13–16^. Joint effects of these exposures are probable and have been seen previously in research on childhood psychopathology^6,17–19^. As exposure to indoor air pollutants and psychosocial factors co-occur and often cluster around socioeconomic status, it is important to consider joint effects of these risk factors. Investigating joint effects will also help to identify and target subgroups that are especially vulnerable to developing childhood psychopathology. However, a limitation of current studies is that they typically focus on exposure at one time period (i.e., exposure during pregnancy). Exposure to both indoor air pollution and psychosocial factors can be reduced through interventions. Understanding the role of these exposures and their joint effects, as well as the timing of exposure, in developing psychopathology will allow for targeted interventions with the goal of reducing incidence and symptoms of childhood psychopathology.

Pregnancy and early life may be particularly important time periods to explore the individual and joint effects of air pollution and psychosocial stress on brain health. The central nervous system begins to develop as early as the first month of gestation^20^ and key milestones in brain development occur throughout pregnancy. After birth, postnatal synaptogenesis, apoptosis, and neuronal pruning further shape the neuronal synapses. Disruption or dysregulation of these processes can lead to functional abnormalities which can lead to psychopathology^21^. Therefore, brain development in pre-and early postnatal time periods may be especially sensitive to indoor air pollutants and psychosocial stressors.

Literature on sensitive periods for the effects of exposure to air pollution on childhood psychopathology are mixed. For example, a German study found that prenatal exposure to environmental tobacco smoke was more strongly associated with behavioral problems at 10 years old^14^, as measured by the Strengths and Difficulties Questionnaire (SDQ),as opposed to early life tobacco exposure. However, a study of French children found postnatal environmental tobacco exposure, alone or in combination with prenatal exposure, was associated with adverse SDQ scores^15^. Additionally, a Chinese study investigating pre-and postnatal exposure to PM_2.5_ and PM_10_ were more strongly associated with adverse neurodevelopment at 2 years old than prenatal exposure^16^.

Few studies have compared how psychosocial factors experienced during pregnancy and early life of the child impact later childhood psychopathology. A systematic review on childhood maltreatment and psychopathology found no consensus on sensitive periods among available epidemiology studies^22^. One study from the United States found prenatal depressive symptoms, as measured by the Edinburgh Postpartum Depression Scale (EPDS), was associated with poorer behavioral development in mid-childhood^23^. This study found postpartum symptoms were not associated with any behavioral outcomes after adjustment for prenatal symptoms^23^. Another study from the United States found prenatal stressful life events and intimate partner violence were associated with externalizing Childhood Behavior Checklist (CBCL) score^24^.

Another limitation of the current literature is that a majority of studies are focused on children in high-income countries (HICs). Pregnant women and children in low-and middle-income countries (LMICs) are particularly impacted by indoor air pollution and psychosocial factors. LMICs typically have higher levels of indoor air pollution, compared to HICs, partially due to fuels used while cooking and heating the home^25^ Women and children are most at risk for exposure to indoor air pollution because they tend to spend more time indoors and are more involved in food preparation and cooking^26^. Compared to HICs and the global average, LMICs also fare worse in social indicators for children, including the human development index, primary school enrollment ratio, under five malnutrition, among others^27^. These social indicators are known to be associated with childhood mental health^27^.

We aimed to address these limitations by leveraging data from the South African Drakenstein Child Health Study (DCHS). Our goal was to examine the individual and joint effects of indoor air pollutants and psychosocial factors in pregnancy and early childhood on childhood psychopathology. This study examines how individual and joint exposure during pregnancy and early life impact childhood psychopathology by investigating each period separately. We additionally use a life course modeling approach to determine sensitive periods for separate exposures. Finally, we investigate sensitive periods of joint exposure effects.

## Methods

### Study Population

Participants came from the Drakenstein Child Health Study (DCHS), a population-based birth cohort from outside Cape Town, South Africa. As previously described in greater detail, pregnant women were recruited between March 2012 and March 2015 from two public sector healthcare clinics. Enrollment criteria were that participants were at least 18 years of age, within 20-28 weeks gestation, and were not planning on moving away from the district. Detailed recruitment information for the DCHS has been described elsewhere^28,29^. The DCHS conducts follow-up visits with mother-child pairs frequently; 6 visits in the first year of life and 6 monthly visits thereafter. The full cohort includes N=1,141 mother-child pairs; we selected a subset for analysis based on availability of indoor air pollution and CBCL measurements (n=599; Figure S1). The DCHS was approved by the Human Research Ethics Committee of the Faculty of Health Sciences, University of Cape Town, by Stellenbosch University and the Western Cape Provincial Research committee. Written informed consent was provided by the mothers for herself and her child and is renewed annually.

### Exposure Assessment

#### Indoor Air Pollution

Indoor air pollution measurements were taken during participants’ 2^nd^ trimester of pregnancy and postnatally at 4 months. As described previously^17,30^, pollutants measured include particulate matter (PM_10_), carbon monoxide (CO), nitrogen dioxide (NO_2_), sulfur dioxide (SO_2_), and Volatile Organic Compounds (VOCs) benzene and toluene. PM_10_ was collected over 24 hours with a personal air sampling pump (SKC AirChek 52®), using a gravimetrically pre-weighted filter. CO was collected over 24 hours using an Altair® carbon monoxide single gas detection unit, electrochemical sensor detection of gas at 10-minute intervals were collected. SO_2_ and NO_2_ were collected over 2 weeks using Radiello® absorbent filters in polyethylene diffusive body. VOCs, including benzene and toluene, were collected over 2 weeks using Markes® thermal desorption tubes^30^. Information on household information including type of home, distance from major road, size of home, number of inhabitants, access to basic amenities, fuels used for cooking and heating, ventilation within homes, and pesticides and cleaning materials used in the home was collected at home visits^30^.

#### Psychosocial Factors and Their Determinants

Psycwwere collected via questionnaire in the 2^nd^ trimester of pregnancy and at 6-10 weeks postnatal, designed to capture multiple dimensions of the psychosocial landscape. *Socioeconomic status* was collected as a sum of indicators of assets owned and utilized by the household (e.g. electricity in the home, etc.). The USDA Household Food Security Scale was used to measure perceived *food insecurity*^31^. *Emotional, physical, and sexual intimate partner violence* (IPV) was assessed using the IPV Questionnaire adapted from the World Health Organization (WHO) multi-country study and the Women’s Health Study in Zimbabwe^32,33^. Only emotional and physical IPV were used in further analyses because few women (<10%) experienced sexual IPV. The World Mental Health Life Events Questionnaire (LEQ) was used to measure *traumatic life experiences and resilience*. Use of *alcohol and tobacco* were assessed using the Alcohol, Smoking, and Substance Involvement Screening Test (ASSIST). Few women (at most 2%) reported use of other substances (e.g., marijuana) and those questionnaires were not included in further analysis. Additionally, tobacco smoke exposure was assessed via urinary cotinine, but only in the prenatal period. The WHO endorsed measure, Self-Reporting Questionnaire (SRQ-20), was used to measure *psychological distress*^34,35^. Finally, the Edinburgh Postnatal Depression Scale (EPDS) was used to measure *depressive symptoms*^36^. These factors were chosen for this study because they have been shown to be risk factors for childhood psychopathology^22–24^.

All exposures were natural-log transformed for linear regression modeling and additionally centered and scaled to have a mean at 0 and standard deviation of 1 before mixture modeling.

### Outcome Assessment

Parent-reported child psychopathology was assessed using the school version of the Child Behavior Checklist (CBCL) at 6.5 years of age^37^. The CBCL, consisting of 113 questions, assesses child behavior using a Likert scale (0 = absent; 1= occurs sometimes; 2 = occurs often) to create a score, defined as total problems score. These questions can be sub divided into internalizing and externalizing sub scales. The internalizing scale includes questions from the anxious/depressed, withdrawn/depressed, and somatic complaints syndromic scales. The externalizing scale includes the rule-breaking and aggressive behavior syndromic scales^37^. CBCL total problems, internalizing problems, and externalizing problems scores were right skewed and were therefore natural log-transformed to be used in modeling approaches described below.

### Statistical Analysis

#### Multiple imputation of missing values

There were no missing values in our outcome or covariates. However, some participants were missing indoor air pollution or psychosocial measurements in either the pre-or postnatal period (Table S1). We assume these values are missing at random based on inspections of the missingness patterns (Figure S2). To increase sample size and retain statistical power we used multiple imputation, as implemented by the *hmisc* R package, to impute missing exposure variables. Pre-and postnatal indoor air pollution and psychosocial factor variables were imputed using predictive mean matching. Separate imputation models were used for each time period and included indoor air pollutants, household characteristics, and psychosocial factor measurements. For each time period, five seed numbers were created using a random number generator. Each seed resulted in its own set (k = 10) of multiple imputed variables, and one imputed set of was randomly chosen to use for analyses Pooling of the k imputed values was not compatible with methods used in the statistical analysis. Finally, the seed with the highest R^2^ values, a measure used to explain how well the missing variable was predicted, was selected for primary analysis and results. Sensitivity analyses using complete cases and the other imputation seeds were conducted.

#### Association analyses

<u>This study aims to investigate both individual and joint effects of indoor air pollutants and psychosocial factors at two time periods, pre-and postnatal, on CBCL score at 6.5 years including externalizing, and internalizing subscales. The analysis workflow consists of two parts, 1) single exposure period exposure-outcome modeling, where pre-and postnatal exposures are investigated separately, and 2) Exposure modeling of sensitive periods. In our single exposure period approach, we first used linear regression to examine individual exposure-outcome effects, and next two environmental mixture methods (Quantile G-Computation and Self-Organizing Maps) to investigate joint effects of multiple exposures (exposure mixture). To investigate sensitive periods of exposure for individual exposures we used a Structured Life Course Modeling Approach and for the exposure mixture we used Self-Organizing Maps as described below (n=599; Figure S1).</u>

#### Pre-and Postnatal exposure-outcome modeling

To investigate the association between each individual prenatal exposure and CBCL scores at 6.5 years we used linear regression. Confounding was assessed using directed acyclic graphs (DAGs) (Figure S3A-C). All models were minimally adjusted for a set of confounders including maternal HIV status, maternal age, ancestry (Black African versus mixed ancestry), and socioeconomic status (when not used as the exposure of interest). In sensitivity analyses an extended confounder set was used to additionally adjust for confounding from psychosocial factors in air pollutant exposure models and vice versa. In the air pollutant exposure models, psychosocial confounders were summarized as principal components (using Principal Components Analysis), as opposed to individual variables, to avoid instability in estimation because psychosocial factors were highly correlated. In these extended confounder set models the first four principal components of psychosocial factors were included as confounders. Similarly, the models investigating socioeconomic status or psychosocial stressors as the main exposures were adjusted for the first four principal components of indoor air pollution.

Two environmental mixture modeling approaches were used to investigate joint effects of the prenatal mixture of indoor air pollutants and psychosocial factors on CBCL: quantile g-Computation, and self-organizing maps (SOM). Quantile g-computation provides an overall mixture effect estimate as well as relative contributions of each exposure in the mixture^38^. Quantile g-computation uses g-computation to estimate the total effect of our mixture as each exposure increases by one quantile simultaneously. Additionally, the partial effect of each exposure is calculated as positive or negative weights^38^. Quantile G-Computation models using the imputed analysis sample data were adjusted for maternal age, maternal HIV status, and ancestry. The adjusted model was fitted for deciles of exposure and 200 bootstrapped samples. All quantile g-computation analyses were conducted using the *qgcomp* package in R (ver. 4.1.2).

SOM was used to create and examine profiles of exposure to indoor air pollution and psychosocial factors on CBCL scores. The SOM algorithm identifies clusters, or profiles, of an exposure mixture. Clusters have exposure levels that are homogenous within the cluster and heterogenous between clusters^39,40^. The number of clusters chosen for further analysis was determined by multiple statistical measures assessing group structure, including Akaike information criterion (AIC), and adjusted R^2^, additionally visual inspection of the clusters for interpretability and appropriate distribution of participants among clusters. To investigate the joint effect of indoor air pollutants and psychosocial factors on CBCL, SOM clusters were then assigned to participants and treated as a categorical exposure variable in linear regression analysis. The linear regression model using SOM cluster as the exposure was adjusted for maternal age, maternal HIV status, and ancestry. We used the SOM R package as implemented in https://github.com/johnlpearce/ECM.

The effect of postnatal exposures to indoor air pollution and psychosocial factors on CBCL at 6.5 years was also assessed using single-exposure linear regression models, Quantile G-Computation, and SOM. Postnatal single-exposure models were conducted and adjusted for confounding in the same way as prenatal single-exposure models, using maternal age and HIV status at baseline, ancestry and SES. Including principal components of the other group of exposures (indoor air pollutants or psychosocial factors) as confounders was also done as a sensitivity analysis. Postnatal mixture modeling (Quantile G-Computation and SOM) was also conducted and adjusted for confounding the same way as prenatal models.

### Sensitive Periods of Exposure

To investigate sensitive periods of exposure to individual indoor air pollutants and psychosocial factors (prenatal vs. postnatal) we used the structured life course modeling approach (SLCMA)^41^. SLCMA is an established approach that can compare multiple competing theoretical models of life course modeling^42–44^. SLCMA uses least angle regression variable selection, a type of least absolute shrinkage and selection operator (LASSO) to determine which a priori determined life course hypothesis is most associated with an outcome. Based on the number of exposure periods and exploratory nature of this study, we investigated four life course hypotheses: sensitive periods (prenatal versus postnatal), accumulation of exposure over the pre-and postnatal periods, and interaction between pre-and postnatal period exposures. All single-exposure SLCMA models were adjusted for maternal age, maternal HIV status, ancestry, and SES (except for when SES was the exposure of interest).

To investigate sensitive periods of the indoor air pollution and psychosocial factor exposure mixture, we used SOM. As opposed to SLCMA which is a structured approach comparing a priori hypotheses, SOM is an unstructured approach that can compare ad hoc hypotheses. As discussed above, SOM creates and compares profiles of exposure mixtures. Using this method in addition to SLCMA enhances our analysis because SOM compares pre-and postnatal exposure profiles experienced by our participants. To create SOM clusters identifying profiles of pre-and postnatal exposure, we added both pre-and postnatal exposures to the SOM algorithm at the same time. These SOM clusters represent combined pre-and postnatal exposure profiles. The SOM clusters were assigned to each participant and used as a categorical exposure variable in linear regression analysis. The linear regression model was adjusted for maternal HIV status and maternal age at baseline, and ancestry.

## Results

### Study Population Characteristics

Among our study population of n=599 participants, the average maternal age was 26.85 years (SD: 5.68), with nearly a quarter of mothers (n=134, 22.4%) HIV-infected at enrolment. Among the children, half were male (n=309, 51.6%), (Table 1). Median IAP exposures were higher in the prenatal period compared to the postnatal period. Median SES was slightly higher and food insecurity was lower in the postnatal period. Other psychosocial factor averages were similar in prenatal and postnatal periods (Table 1). Indoor air pollutants were not highly correlated within or between time periods; psychosocial factors were moderately correlated within and between time periods (Figure S4). At 6.5 years of age, the CBCL total problems score was highly correlated with both externalizing (Pearson rho = 0.91) and internalizing (Pearson Rho = 0.73) sub scales. Externalizing and internalizing sub scales were moderately correlated with each other (Pearson Rho = 0.48; Table S2).

**Table 1.**
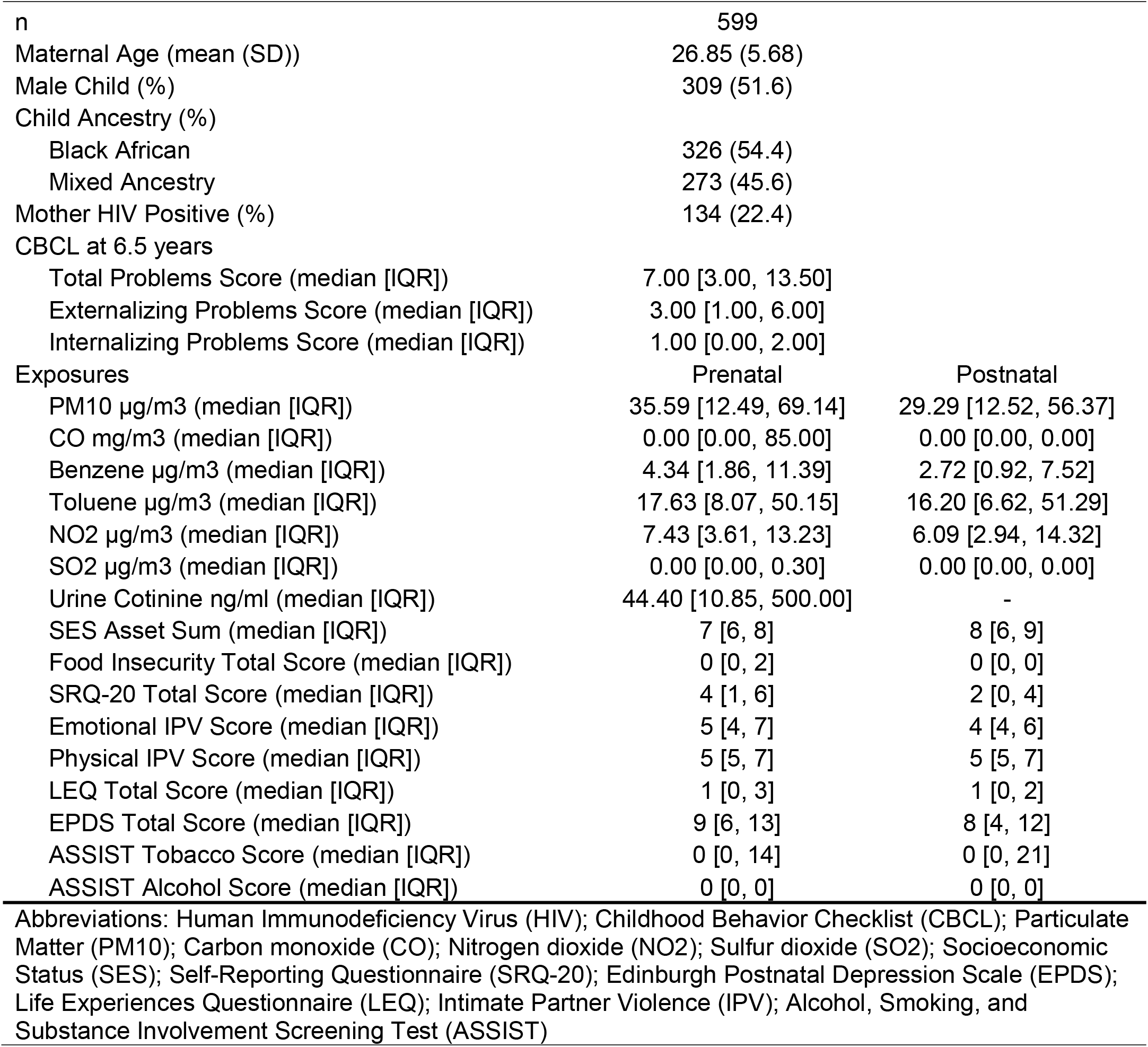
Drakenstein Child Health Study (DCHS) population characteristics.

### Prenatal Exposure Analyses

In adjusted single-exposure linear regression models for the prenatal exposures a one-unit increase in log-transformed PM_10_ (beta: 0.08; 95% CI: 0.03, 0.13), psychological distress (SRQ) (beta: 0.10; 95% CI: 0.00, 0.20), Physical IPV (beta: 0.29; 95% CI: 0.03, 0.55), depression (EPDS) (beta: 0.16; 95% CI: 0.04, 0.28), and ASSIST Alcohol (beta: 0.11; 95% CI: 0.03, 0.19) were associated with an increase in the CBCL total problems score (Table S3, Figure 1A). Adjusted single-exposure models investigating CBCL externalizing problems found associations between a one-unit increase in log-transformed prenatal Toluene (beta: 0.05; 95% CI: 0.00, 0.10), and ASSIST Alcohol (beta:0.11; 95% CI: 0.04, 0.18; Table S3, Figure 1C). For CBCL internalizing problems, we found associations with a one-unit increase in log-transformed prenatal PM10 (beta: 0.04; 95% CI: 0.00, 0.08), SRQ (beta: 0.09; 95% CI: 0.02, 0.17), and

**Figure 1.**
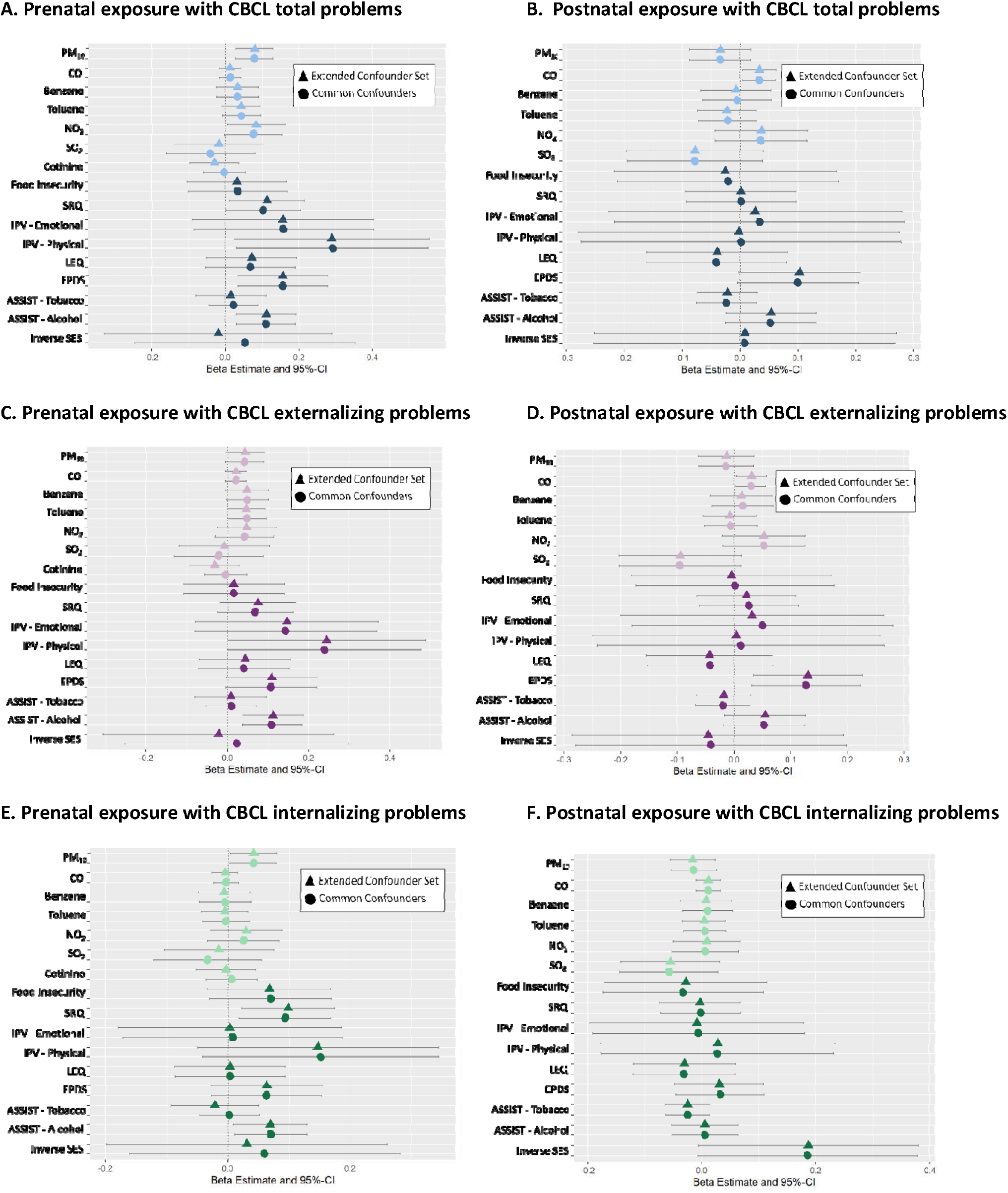
Results from single-exposure linear regression models. Common confounder models adjusted for maternal age, maternal HIV status, ancestry, and SES (when SES is not the main exposure). In extended confounder models, indoor air pollutant models are additionally adjusted for principal components of psychosocial exposures, and vice versa. A. Associations between prenatal exposures and CBCL total problems. B. Associations between postnatal exposures and CBCL total problems. C. Associations between prenatal exposures and CBCL externalizing problems. D. Associations between postnatal exposures and CBCL externalizing problems. E. Associations between prenatal exposures and CBCL internalizing problems. F. Associations between postnatal exposures and CBCL internalizing problems. A. Prenatal exposure with CBCL total problems. B. Prenatal exposure with CBCL externalizing problems. C. Prenatal exposure with CBCL internalizing problems D. Postnatal exposure with CBCL total problems. E. Postnatal exposure with CBCL externalizing problems. F. Postnatal exposure with CBCL internalizing problems

ASSIST Alcohol (beta: 0.07; 95% CI: 0.01, 0.13; Table S3, Figure 1E). Results were similar in the complete cases analyses and when using other imputation seeds (Table S3). Results were similar when single-exposure linear regression models were additionally adjusted for PCs of the other exposure group (IAP or PF). The only difference between the two adjustment sets was that in the extended models, NO_2_ was associated with CBCL total problems and physical IPV was associated with CBCL externalizing problems.

Adjusted quantile g-computation models showed that the prenatal mixture was significantly associated with higher CBCL total scores (Psi beta: 0.32; p-value: 0.013) and externalizing (Psi beta: 0.35; p-value: 0.003) problems, and not with internalizing (Psi beta: 0.14; p-value: 0.19) problems (Figure 2A-C). Prenatal ASSIST Alcohol score had the largest partial effect in both total and externalizing problems models (Figure S5A-B).

**Figure 2.**
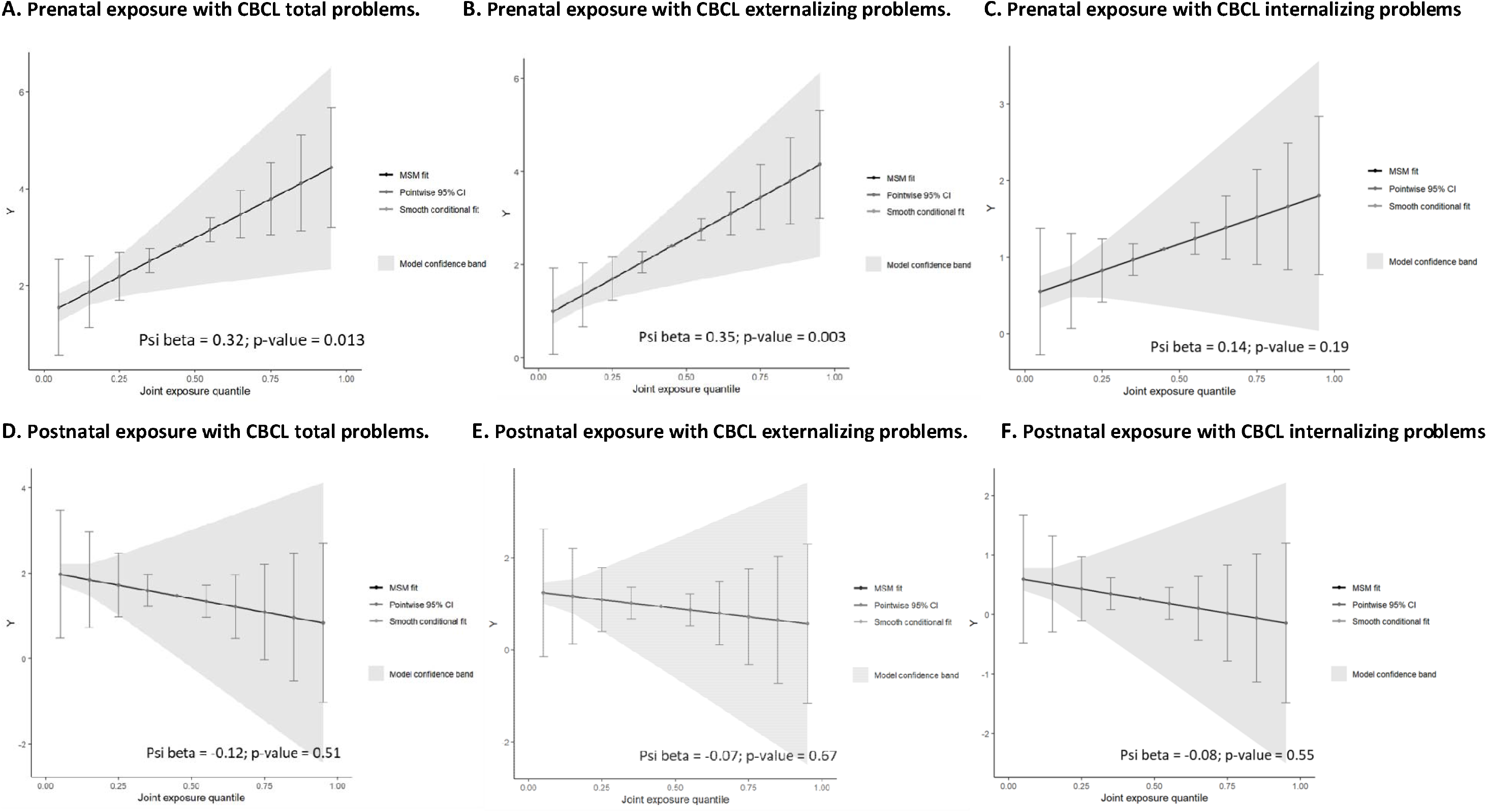
Total mixture effect estimates from quantile G-computation models, adjusted for maternal age, maternal HIV status, and ancestry. A. association between prenatal exposure and CBCL total problems. B. Association between prenatal exposure mixture and CBCL externalizing problems. C. Association between prenatal exposure mixture and CBCL internalizing problems. D. Association between postnatal exposure mixture and CBCL total problems. E. Association between prenatal exposure mixture and CBCL externalizing problems. F. Association between postnatal exposure mixture and CBCL internalizing problems.

SOM analysis using prenatal exposures grouped our study population in 4 exposure profile clusters. Cluster 3 was selected as the reference cluster because it had the lowest exposure to all pollutants and psychosocial factors, except for SO2 (Table S7, Figure S6A). Using SOM cluster indicator as categorical exposure in adjusted linear regression model, cluster 2 (beta: 0.24; 95% CI: 0.10, 0.58), which was reflective of smoking and alcohol exposures (high cotinine level and ASSIST Tobacco and Alcohol scores) was associated with increasing CBCL total problems score, compared to the reference cluster 3. Additionally, cluster 4 (beta: 0.31; 95% CI: 0.07, 0.55), the cluster reflective of high levels of most psychosocial factors, was associated with increasing CBCL total problems score, compared to the reference cluster 3 (Table S8, Figure S6B). Similarly, cluster 2 (beta: 0.27; 95% CI: 0.05, 0.50) and cluster 4 (beta: 0.26; 95% CI: 0.04, 0.48) were associated with increasing CBCL externalizing problems score, compared to the reference cluster 3 (Table S8, Figure S6C). No SOM cluster was significantly associated with CBCL internalizing problems, though all were negatively associated compared to cluster 3 (Table S8, Figure S5D).

### Postnatal Exposure Analyses

In adjusted single-exposure models for the postnatal exposures, only a one-unit increase in log-transformed CO (beta: 0.03; 95% CI: 0.00, 0.06) was associated with CBCL total problems (Table S4, Figure 1B). A one-unit increase in log-transformed CO (beta: 0.03; 95% CI: 0.00, 0.06) and EPDS (beta: 0.13; 95% CI: 0.03, 0.23) were associated with CBCL externalizing problems (Table S4, Figure 1D). No postnatal exposures were significantly associated with CBCL internalizing problems in adjusted single-exposure models (Table S4, Figure 1F). Results were similar in the complete cases analyses and when using other imputation seeds (Table S4).

The postnatal exposure mixture was not significantly associated with any CBCL outcome in quantile g-computation model (Figure 2D-F, Figure S5D-F).

SOM analysis using postnatal exposures grouped our study population in 4 exposure profile clusters, which differed from prenatal clusters (Table S9, Figure S7A). For linear regression modeling, cluster 3 was again used as the reference group as it had the lowest exposure medians for all exposures (Table S9). No postnatal exposure cluster, compared to the reference cluster 3, was significantly associated with any CBCL outcome (Table S10, Figure S7B-D).

### Sensitive Period Analyses

Single-exposure SLCMA analysis for CBCL total problems score confirmed the prenatal sensitive period as the most likely life course model for PM10, psychological distress (SRQ) and Physical IPV, and the postnatal sensitive period for CO. However, an accumulation life course model was most supported for depression (EPDS) and ASSIST Alcohol. Additionally, an interaction life course model was most supported for Emotional IPV. Similar patterns were seen for CBCL internalizing and externalizing problems (Table 2).

**Table 2.** Results from single-exposure structured life course modeling approach (SLCMA) analyses. Single-exposure SLCMA models adjusted for maternal age, maternal HIV status, ancestry, and socioeconomic status (when not the exposure of interest).

| Total Problems |  |  |  |  |  |
| --- | --- | --- | --- | --- | --- |
| Relaxed LASSO Inference |  |  |  |  |  |
| Exposure | 1st selection in LASSO | R2 | Coef | CI low | CI up |
| **PM10** | **Prenatal** | **0.016** | **0.123** | **0.044** | **0.201** |
| **CO** | **Postnatal** | **0.008** | **0.089** | **0.011** | **0.168** |
| Benzene | Interaction | 0.005 | -0.066 | -0.142 | 0.010 |
| Toluene | Prenatal | 0.005 | 0.069 | -0.010 | 0.147 |
| NO2 | Accumulation | 0.006 | 0.054 | 0.000 | 0.108 |
| SO2 | Accumulation | 0.003 | -0.038 | -0.093 | 0.017 |
| SES Inverse | Prenatal | 0.001 | -0.191 | -0.664 | 0.256 |
| Food Insecurity | Prenatal | 0.000 | 0.022 | -0.065 | 0.110 |
| **SRQ** | **Prenatal** | **0.007** | **0.082** | **0.002** | **0.162** |
| **IPV Emotional** | **Interaction** | **0.009** | **-0.069** | **-0.128** | **-0.012** |
| **IPV Physical** | **Prenatal** | **0.008** | **0.089** | **0.010** | **0.168** |
| LEQ | Interaction | 0.002 | -0.044 | -0.121 | 0.033 |
| **EPDS** | **Accumulation** | **0.012** | **0.080** | **0.021** | **0.140** |
| ASSIST Tobacco | Postnatal | 0.001 | -0.035 | -0.113 | 0.044 |
| **ASSIST Alcohol** | **Accumulation** | **0.013** | **0.076** | **0.023** | **0.131** |
| Externalizing Problems |  |  |  |  |  |
| Relaxed LASSO Inference |  |  |  |  |  |
| Exposure | 1st selection in LASSO | R2 | Coef | CI low | CI up |
| PM10 | Prenatal | 0.005 | 0.064 | -0.008 | 0.136 |
| **CO** | **Accumulation** | **0.01** | **0.06** | **0.013** | **0.108** |
| Benzene | Prenatal | 0.006 | 0.068 | -0.005 | 0.14 |
| **Toluene** | **Prenatal** | **0.007** | **0.074** | **0.002** | **0.146** |
| NO2 | Accumulation | 0.005 | 0.0449 | -0.005 | 0.095 |
| SO2 | Postnatal | 0.005 | -0.065 | -0.137 | 0.008 |
| SES Inverse | Interaction | 0.002 | -0.031 | -0.092 | 0.03 |
| Food Insecurity | Prenatal | 0 | 0.011 | -0.07 | 0.092 |
| **SRQ** | **Prenatal** | **0.004** | **0.054** | **-0.019** | **0.128** |
| **IPV Emotional** | **Interaction** | **0.007** | **-0.055** | **-0.108** | **-0.001** |
| **IPV Physical** | **Prenatal** | **0.006** | **0.073** | **0.0002** | **0.145** |
| LEQ | Postnatal | 0.001 | -0.028 | -0.102 | 0.046 |
| **EPDS** | **Accumulation** | **0.014** | **0.0799** | **0.023** | **0.134** |
| ASSIST Tobacco | Interaction | 0.004 | 0.058 | -0.0136 | 0.13 |
| **ASSIST Alcohol** | **Accumulation** | **0.015** | **0.076** | **0.0256** | **0.126** |
| Internalizing Problems |  |  |  |  |  |
| Relaxed LASSO Inference |  |  |  |  |  |
| Exposure | 1st selection in LASSO | R2 | Coef | CI low | CI up |
| **PM10** | **Prenatal** | **0.008** | **0.0645** | **0.0064** | **0.122** |
| CO | Postnatal | 0.002 | 0.0321 | -0.026 | 0.09 |
| Benzene | Interaction | 0.002 | -0.028 | -0.084 | 0.029 |
| Toluene | Interaction | 0 | 0.0158 | -0.042 | 0.074 |
| NO2 | Prenatal | 0.001 | 0.0256 | -0.032 | 0.084 |
| SO2 | Accumulation | 0.003 | -0.029 | -0.0699 | 0.011 |
| SES Inverse | Postnatal | 0.005 | 0.057 | -0.055 | 0.119 |
| Food Insecurity | Prenatal | 0.003 | 0.0459 | -0.019 | 0.111 |
| **SRQ** | **Prenatal** | **0.1** | **0.074** | **0.015** | **0.133** |
| **IPV Emotional** | **Interaction** | **0.013** | **-0.062** | **-0.105** | **-0.019** |
| IPV Physical | Prenatal | 0.004 | 0.0462 | -0.0123 | 0.105 |
| LEQ | Interaction | 0.007 | -0.058 | -0.115 | -0.001 |
| EPDS | Prenatal | 0.003 | 0.063 | -0.026 | 0.154 |
| ASSIST Tobacco | Postnatal | 0.003 | -0.037 | -0.095 | 0.021 |
| **ASSIST Alcohol** | **Prenatal** | **0.009** | **0.072** | **0.011** | **0.132** |

Footnotes: Relaxed LASSO results are not adjusted for multiple testing nor for lasso selection. Bolded values indicate statistical significance.
Abbreviations: Human Immunodeficiency Virus (HIV); Childhood Behavior Checklist (CBCL); Particulate Matter (PM10); Carbon monoxide (CO); Nitrogen dioxide (NO2); Sulfur dioxide (SO2); Socioeconomic Status (SES); Self-Reporting Questionnaire (SRQ-20); Edinburgh Postnatal Depression Scale (EPDS); Life Experiences Questionnaire (LEQ); Intimate Partner Violence (IPV); Alcohol, Smoking, and Substance Involvement Screening Test (ASSIST)

SOM analysis with both prenatal and postnatal exposures grouped our study population in 6 exposure profile clusters. Custer 6 was selected as the reference group for regression analyses because it has the lowest median exposure values for most exposures (Table S5, Figure 3A). Adjusted linear regression models using SOM cluster as the exposure found cluster 1 (beta: 0.38; 95% CI: 0.14, 0.62), characterized by low SES, high EPDS and food insecurity scores in both pre-and postnatal periods as well as high prenatal indoor air pollution, was significantly associated with CBCL total problems, compared to the reference cluster 6 (Table S6, Figure 3B). Cluster 1 (beta: 0.29; 95% CI: 0.07, 0.51) was also associated with CBCL externalizing problems, compared to the reference cluster 6 (Table S6, Figure 3C). No SOM cluster was associated with CBCL internalizing problems score (Table S6, Figure 3D).

**Figure 3.**
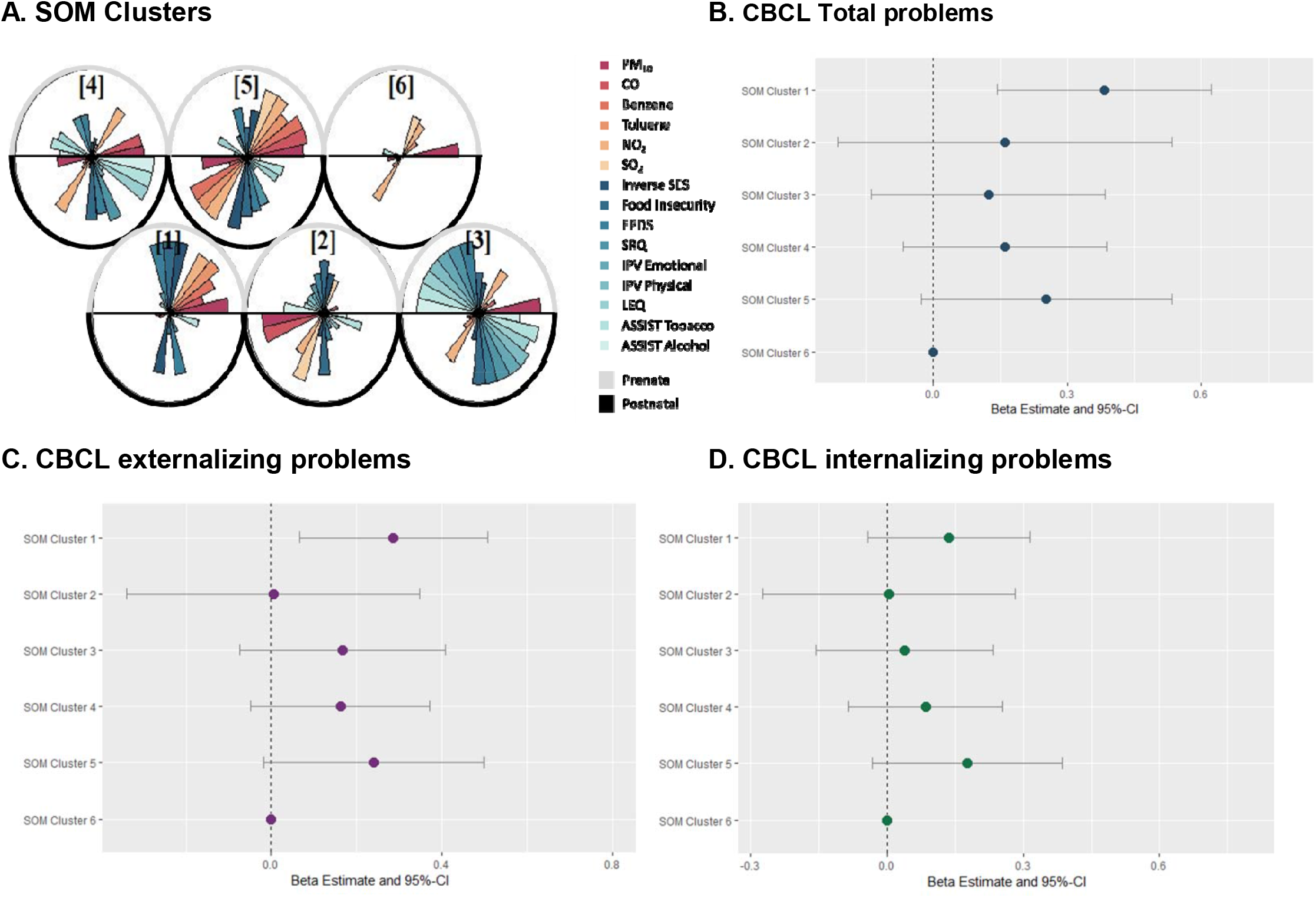
Results from self-organizing map (SOM) analysis combining pre- and postnatal indoor air pollutants and psychosocial factors. A. SOM clusters created using pre-and postnatal indoor air pollutants and psychosocial factors. B. Associations between SOM clusters and CBCL total problems, adjusted by maternal age, maternal HIV status, and ancestry. SOM cluster 6 is used as the reference group. C. Associations between SOM clusters and CBCL externalizing problems, adjusted by maternal age, maternal HIV status, and ancestry. SOM cluster 6 is used as the reference group. D. Associations between SOM clusters and CBCL internalizing problems, adjusted by maternal age, maternal HIV status, and ancestry. SOM cluster 6 is used as the reference group.

## Discussion

This study of mother-child pairs in the South African Drakenstein Child Health Study found prenatal exposure to indoor air pollution, as opposed to postnatal, was most strongly associated with childhood psychopathology at 6.5 years. This result was seen in both traditional single-exposure, single-period modeling, as well as in methods using environmental mixture and structured life course approaches. Only CO in the postnatal period had stronger associations with child psychopathology, as compared to the prenatal period. We also found exposure to psychosocial factors including depression, ASSIST Alcohol score, and psychological distress in the prenatal period to be most strongly associated with child psychopathology. This was observed in single-exposure, single-time period models as well as single-time period environmental mixtures modeling approaches Additionally, the SOM model combining pre-and postnatal exposures found the cluster with high depression scores in both periods, along with high prenatal indoor air pollution, was associated with CBCL scores.

PM_10_ exposure during pregnancy was found to be especially harmful to childhood psychopathology at age 6.5 years. This extends our previous work in the DCHS, in which indoor PM_10_ exposure in the prenatal period was also associated with impaired neurodevelopment at 2 years, by showing adverse associations with cognitive outcomes not only in early childhood but also at school-age^45^. A systematic review of prenatal exposure to outdoor PM_10_ and psychopathology, like autism spectrum disorder (ASD) or attention deficit and hyperactivity disorder (ADHD), found inconsistent results which may be due to differential exposure and differing outcome assessments between studies^46^. Further research is necessary to elucidate the association of prenatal PM_10_ exposure and psychopathology. When investigating sensitive periods of exposure, a Chinese study investigating ambient PM exposure during pregnancy and the first 2 years postnatal found both pre-and postnatal exposure to PM_10_ and PM_2.5_ were associated with adverse Chinese revision of Bayley Scale of Infant Development (BSID-CR) scores. However, in that study postnatal exposure was more strongly associated with BSID-CR scores than prenatal exposure^16^, which is the opposite of our findings. Differences in findings could be due to differences in exposure measurement and concentrations. In the Chinese study, authors investigated ambient PM exposure as opposed to indoor PM, and the exposure concentrations were much higher in the Chinese study. Their minimum ambient prenatal PM_10_ concentration was 97.93 μg/m^3^, which is much larger than our median indoor PM_10_ concentration of 35.59 μg/m^3^. They also investigated postnatal PM_10_ exposure for 2 years after birth, compared to 4 months in our study^16^. Another study investigating sensitive periods of air pollution exposure and cognitive functioning found ambient PM_10_ exposure in the 22^nd^-29^th^ weeks of pregnancy, and not postnatal exposure, was associated with cognitive function in boys at 5-6 years old^47^. However, this study used postnatal exposure at 60 months, which is much later than in our study^47^. Most research on air pollution exposure focuses on one single time period, as shown in a recent review^48^. Additionally there are few epidemiological studies that investigate early life or childhood exposure to air pollution and childhood psychopathology^48^. Most studies explore exposure and psychopathology in adulthood, or only prenatal exposure on childhood psychopathology^48^.

Postnatal, but not prenatal, CO exposure was associated with childhood psychopathology at 6.5 years old. There are very few studies examining the effect of pre-and postnatal CO exposure on child psychopathology. One study by Dix-Cooper, et al. found CO exposure during the 3^rd^ trimester from woodsmoke to be associated with decreased performance on several neurobehavioral tests at 6-7 years old. Exposure to CO as an infant (0-9 months) was not associated with neurobehavioral tests at 6-7 years^49^. Differential findings could be due to low sample size (n = 20 in prenatal, n = 39 in postnatal) in their study, as well as measurement of CO exposure and neurobehavioral outcome measures. Their study used the average of personal (wearable) measurement devices, while our study measured CO in the main living area of the home^49^. Based on these findings, there is a need for more research on how CO exposure in early life and childhood impacts childhood psychopathology.

Our study found prenatal maternal smoking was associated with childhood psychopathology,. Maternal tobacco use during pregnancy is well known to be associated with decreased neurodevelopment and child behavior problems^50^ One hypothesized pathway linking prenatal tobacco use and neurodevelopment and psychopathology is through low birthweight and decreased in-utero brain growth^51^. In support of our findings, prenatal tobacco use has been seen to affect inattention, oppositional behavior, emotional instability, and physical aggression^50^. In a previous study done in the DCHS prenatal, but not postnatal, tobacco use was associated with decreased adaptive behavior^45^. Studies investigating environmental tobacco smoke (ETS) exposure during the pre-and postnatal periods have mixed findings regarding sensitive periods. A German study investigating ETS exposure during pregnancy and early life found prenatal exposure was more strongly associated with SDQ score at 10 years old^14^ than postnatal exposure, which is in line with our findings. However, a French study found postnatal ETS exposure, alone or associated with prenatal exposure, was associated with adverse SDQ scores^15^. Additional research is needed to fully clarify sensitive periods of ETS exposure.

We also found associations between pre-and postnatal maternal alcohol and childhood psychopathology. Single-exposure, single-period models found associations between ASSIST Alcohol score and psychopathology in the prenatal period, but not in the postnatal period. However, SLCMA modeling found accumulation of ASSIST alcohol score in the pre-and postnatal periods impacted psychopathology. These results indicate that alcohol exposure during pregnancy is more strongly associated with childhood psychopathology, but that maternal drinking behaviors postnatally may also contribute to psychopathology. Alcohol is a known teratogen that disrupts fetal development. Fetal alcohol spectrum disorder (FASD) has a wide range of both physical and neurological symptoms including, growth deficiency, abnormal brain growth, and cognitive and behavioral impairments^52^. The teratogenic effects of alcohol on the developing fetal brain may explain the stronger association with prenatal exposure and CBCL score in single-time period analyses in this study. However, a systematic review found mixed associations between alcohol consumption during pregnancy and childhood ADHD incidence, an externalizing psychopathology^53^. While alcohol use after pregnancy is not neurotoxic to the child in the same way as to the fetus, maternal alcohol use may impact the child through other pathways, such as parenting skills.

Most psychosocial factors investigated were not associated with childhood psychopathology in either time period. However, pre-and postnatal depression was also associated with childhood psychopathology. The relationship between maternal depression and child psychopathology has been well established, and maternal depression is a distinct early life stress for the child that shapes the child’s stress response^54^. Results from a birth cohort measuring the effects of postnatal depression in mothers on immune markers and child psychopathology found that children of depressed mothers exhibited higher immune markers and greater social withdrawal. They also found that depressed mothers had higher cortisol and immune makers, and displayed more negative parenting behaviors^54^. It is difficult to investigate sensitive periods of exposure to psychosocial factors, like depression, because unlike with pollutants these factors may vary less over this small time interval. Consequently, one major limitation of this analysis is the lack of participants with low psychosocial factor exposure during one period, and high exposure in another period. In our study, pre-and postnatal measurements were taken less than 1 year apart and exposure to these factors did not change very much between time periods. This limits comparisons of sensitive periods of exposure, as we cannot compare high/low and low/high groups.

Joint effects of indoor air pollution and psychosocial factors occurred, primarily in the prenatal period. Quantile g-computation analysis found increases in the total prenatal exposure mixture were associated with increased CBCL scores. This is in line with our previous in the DCHS, which found joint effects of prenatal exposure to indoor air pollution and psychosocial factors on trajectories of childhood psychopathology from 2 to 5 years old ^17^. Other studies using interaction terms between individual pollutants and psychosocial factors instead of environmental mixture methodology, have also found joint effects of environmental and psychosocial exposures on childhood psychopathology^6,18^. A New York City, USA, based study found interactions between prenatal polycyclic aromatic hydrocarbons (PAH) and early life stress on CBCL attention and thought scores^6^. That same cohort also found interactions between prenatal PAH exposure and prenatal/childhood hardship on ADHD behavior problems^18^. Synergy between indoor air pollutants and psychosocial factors may derive from mechanisms involving inflammation and oxidative stress^9,55^. Exposure to both air pollution^9,56–59^ and stress associated with psychosocial factors^55,59,60^ have been associated with inflammation and oxidative stress. Inflammation and oxidative stress can damage neurons and the CNS, which may impact psychopathology later in life^9^. Future studies should investigate these factors as mediators between exposure to air pollution and psychosocial factors and childhood psychopathology.

This study has several limitations. First, exposure measurements for fine (PM_2.5_) and ultrafine PM measurements were not collected in this cohort because, at the time, personal PM_2.5_ monitoring was not easily available for a study of this size. PM smaller than PM_10_ is hypothesized to be more harmful to the CNS because its smaller size allows for particles to travel throughout the body and brain^9^. Additionally, each air pollutant measurement was only collected once in each time period. There could be some misclassification of the exposure, though prior DCHS studies have also found associations between indoor air pollutants and several neurodevelopmental and psychopathological outcomes^17,45^, as well as respiratory outcomes^61^. Another small limitation in our study is the lack of postnatal maternal cotinine measurements. In the prenatal period our study used urine cotinine measurements, which measure exposure to nicotine from both first and secondhand smoking. Maternal cotinine measurement was not available in the postnatal period, so we could not use it as a measure of maternal smoking. Child cotinine measurements were available in this period, though those would reflect exposure to tobacco smoke from anyone in the household and would not be in concordance with maternal ASSIST Tobacco measurements. Additionally, our methods did not account for variation due to imputation because we were unable to pool imputed datasets. There are also limitations regarding biases from residual confounding, selection bias and generalizability. The DCHS cohort was created to be population based and representative of peri-urban populations in South Africa, and other LMICs, which decreases the likelihood of selection bias. However, these results may not be generalizable to populations outside of peri-urban populations in LMICs.

There are also several strengths of this study worth highlighting. First, the DCHS is a unique multidisciplinary cohort from an underrepresented population in mental health research, children from a LMIC. The DCHS also has repeated measures of indoor air pollutants and psychosocial factor exposures, allowing for comparison of time-period effects on childhood psychopathology and estimation of joint effects of both exposure groups. This cohort also has measures from many domains of psychosocial factors including, threat and trauma, deprivation, substance use, and psychological distress and psychiatric disorders, which allows for a well-rounded view of psychosocial factors and their determinants. This rich dataset allowed us to use both traditional single-exposure linear regression analyses as well as newer environmental mixture methods such as quantile g-computation, SOM, and ECM to examine joint effects of exposures. Additionally, the repeated exposure measurements allowed for testing of sensitive periods of both individual and joint exposure effects using SLCMA and SOM.

This study identified the prenatal period as a particularly vulnerable period for exposure to indoor air pollution and psychosocial factors and later childhood psychopathology. Prenatal exposure to indoor air pollution was seen to be more strongly associated in the prenatal period, while psychosocial factors such as depression and alcohol use were associated with psychopathology in both periods, but had stronger associations in the prenatal period. Interventions aimed at reducing incidence and symptoms of childhood psychopathology should focus on reducing exposure from air pollutants and smoking particularly in the prenatal period. Interventions and mental health services for mother with depression and substance use could be beneficial in both the pre-and postnatal periods. Future studies should investigate additional pollutants such as fine and ultrafine PM, sensitive time periods during pregnancy, and interventions for reducing exposure to indoor air pollutants and psychosocial factors.

## Data Availability

All data produced in the present study are available upon reasonable request to the authors

