## Supplemental Materials for "Sensitive periods for exposure to indoor air pollutants and psychosocial factors in association with symptoms of psychopathology at school-age in a South African birth cohort"

**Tables**

**Table S1**. Proportion of missing pre- and postnatal exposure data in analysis sample.

|  | Prenatal Measurement | | | | Postnatal Measurement | | | |
| --- | --- | --- | --- | --- | --- | --- | --- | --- |
| **Exposure** | **# missing** | **% missing** | **n** | **total N** | **# missing** | **% missing** | **n** | **total N** |
| PM10 | 59 | 10% | 540 | 599 | 253 | 42% | 346 | 599 |
| CO | 115 | 19% | 484 | 599 | 292 | 49% | 307 | 599 |
| benzene | 67 | 11% | 532 | 599 | 285 | 48% | 314 | 599 |
| toluene | 67 | 11% | 532 | 599 | 285 | 48% | 314 | 599 |
| NO2 | 64 | 11% | 535 | 599 | 238 | 40% | 361 | 599 |
| So2 | 64 | 11% | 535 | 599 | 236 | 39% | 363 | 599 |
| maternal smoking (cotinine)* | 17 | 3% | 582 | 599 | - | - | - | - |
| food insecurity | 49 | 8% | 550 | 599 | 119 | 20% | 480 | 599 |
| SRQ | 54 | 9% | 545 | 599 | 197 | 33% | 402 | 599 |
| IPV - emotional | 53 | 9% | 546 | 599 | 196 | 33% | 403 | 599 |
| IPV - physical | 53 | 9% | 546 | 599 | 196 | 33% | 403 | 599 |
| LEQ | 67 | 11% | 532 | 599 | 198 | 33% | 401 | 599 |
| EPDS | 54 | 9% | 545 | 599 | 197 | 33% | 402 | 599 |
| ASSIST - tobacco | 58 | 10% | 541 | 599 | 196 | 33% | 403 | 599 |
| ASSIST - alcohol | 58 | 10% | 541 | 599 | 196 | 33% | 403 | 599 |
| SES assets | 0 | 0% | 599 | 599 | 122 | 20% | 477 | 599 |
| *Cotinine was only measured in the prenatal period | | | |  |  |  |  |  |
| Abbreviations: Particulate Matter (PM10); Carbon monoxide (CO); Nitrogen dioxide (NO2); Sulfur dioxide (SO2); Socioeconomic Status (SES); Self-Reporting Questionnaire (SRQ-20); Edinburgh Postnatal Depression Scale (EPDS); Life Experiences Questionnaire (LEQ); Intimate Partner Violence (IPV); Alcohol, Smoking, and Substance Involvement Screening Test (ASSIST) | | | | | | | | |

**Table S2**. Pearson Correlation between CBCL sub-scores at 6.5 years old.

|  | Total Problems | Externalizing Problems | Internalizing Problems |
| --- | --- | --- | --- |
| Total Problems | 1 | 0.91 | 0.73 |
| Externalizing Problems | 0.91 | 1 | 0.48 |
| Internalizing Problems | 0.73 | 0.48 | 1 |

**Table S3**. Beta estimates and 95% CIs for individual prenatal exposure adjusted linear regression models. The common confounder linear regression models were adjusted for maternal HIV status, maternal age, ancestry, and socioeconomic status. Extended confounder set models using indoor air pollutant exposures were additionally adjusted principal components of psychosocial factors, and vice versa. Tables shows results from complete case models as well as multiple imputation (MI) models using 5 different random seeds (MI1 to MI5). MI5 models were presented in the main analysis.

|  |  | Complete Case | MI1 | MI2 | MI3 | MI4 | MI5 (Analysis Sample) |
| --- | --- | --- | --- | --- | --- | --- | --- |
|  |  | CBCL Total Problems | | | | | |
| PM10 | Extended Confounder Set | **0.08 (0.01, 0.16)** | **0.06 (0.01, 0.11)** | **0.06 (0.01, 0.11)** | **0.06 (0.01, 0.11)** | 0.05 (0, 0.1) | **0.08 (0.03, 0.13)** |
|  | Common Confounders | **0.08 (0.01, 0.16)** | **0.06 (0.01, 0.12)** | **0.06 (0.01, 0.11)** | **0.06 (0.01, 0.12)** | 0.05 (0, 0.11) | **0.08 (0.03, 0.13)** |
| CO | Extended Confounder Set | 0.01 (-0.03, 0.05) | 0.01 (-0.02, 0.04) | 0.01 (-0.02, 0.04) | 0.01 (-0.02, 0.04) | 0.02 (-0.01, 0.04) | 0.01 (-0.01, 0.04) |
|  | Common Confounders | 0.01 (-0.02, 0.05) | 0.01 (-0.02, 0.04) | 0.01 (-0.02, 0.03) | 0.01 (-0.02, 0.04) | 0.02 (-0.01, 0.05) | 0.01 (-0.01, 0.04) |
| Benzene | Extended Confounder Set | 0.03 (-0.05, 0.11) | 0.03 (-0.03, 0.09) | 0.03 (-0.03, 0.08) | 0.04 (-0.02, 0.09) | 0.04 (-0.02, 0.1) | 0.03 (-0.02, 0.09) |
|  | Common Confounders | 0.03 (-0.04, 0.11) | 0.03 (-0.03, 0.09) | 0.03 (-0.03, 0.08) | 0.03 (-0.02, 0.09) | 0.03 (-0.02, 0.09) | 0.03 (-0.02, 0.09) |
| Toluene | Extended Confounder Set | 0.04 (-0.02, 0.11) | 0.03 (-0.02, 0.08) | 0.03 (-0.02, 0.08) | 0.04 (-0.01, 0.09) | 0.02 (-0.03, 0.08) | 0.04 (-0.01, 0.1) |
|  | Common Confounders | 0.04 (-0.02, 0.11) | 0.03 (-0.02, 0.08) | 0.03 (-0.02, 0.08) | 0.04 (-0.02, 0.09) | 0.02 (-0.03, 0.07) | 0.04 (-0.01, 0.1) |
| NO2 | Extended Confounder Set | **0.11 (0.01, 0.22)** | **0.09 (0.01, 0.17)** | 0.07 (-0.01, 0.14) | **0.08 (0, 0.16)** | **0.09 (0.02, 0.17)** | **0.08 (0.01, 0.16)** |
|  | Common Confounders | 0.1 (-0.01, 0.2) | 0.08 (0, 0.15) | 0.06 (-0.02, 0.14) | 0.07 (-0.01, 0.15) | **0.09 (0.01, 0.16)** | 0.08 (0, 0.16) |
| SO2 | Extended Confounder Set | 0.12 (-0.05, 0.29) | -0.05 (-0.15, 0.06) | -0.05 (-0.16, 0.07) | 0 (-0.13, 0.12) | -0.03 (-0.15, 0.09) | -0.02 (-0.14, 0.1) |
|  | Common Confounders | 0.09 (-0.07, 0.26) | -0.07 (-0.18, 0.04) | -0.07 (-0.18, 0.04) | -0.03 (-0.15, 0.09) | -0.05 (-0.17, 0.06) | -0.04 (-0.16, 0.08) |
| Cotinine | Extended Confounder Set | -0.1 (-0.2, -0.01) | -0.02 (-0.08, 0.04) | -0.03 (-0.09, 0.04) | -0.02 (-0.08, 0.05) | -0.03 (-0.09, 0.04) | -0.03 (-0.09, 0.04) |
|  | Common Confounders | -0.08 (-0.16, -0.01) | 0 (-0.06, 0.06) | 0 (-0.06, 0.06) | 0 (-0.06, 0.06) | 0 (-0.06, 0.05) | 0 (-0.06, 0.06) |
| Food Insecurity | Extended Confounder Set | -0.03 (-0.22, 0.15) | 0.07 (-0.07, 0.21) | 0.04 (-0.09, 0.18) | 0.02 (-0.11, 0.16) | 0.05 (-0.08, 0.18) | 0.03 (-0.1, 0.17) |
|  | Common Confounders | -0.05 (-0.24, 0.13) | 0.07 (-0.06, 0.21) | 0.05 (-0.09, 0.18) | 0.03 (-0.11, 0.16) | 0.05 (-0.08, 0.18) | 0.03 (-0.1, 0.17) |
| SRQ | Extended Confounder Set | **0.14 (0, 0.28)** | **0.14 (0.04, 0.24)** | **0.12 (0.02, 0.22)** | **0.13 (0.03, 0.23)** | **0.13 (0.03, 0.23)** | **0.11 (0.01, 0.22)** |
|  | Common Confounders | 0.12 (-0.02, 0.25) | **0.13 (0.03, 0.23)** | **0.11 (0.01, 0.21)** | **0.12 (0.02, 0.22)** | **0.12 (0.02, 0.22)** | **0.1 (0, 0.2)** |
| IPV Emotional | Extended Confounder Set | 0.11 (-0.21, 0.44) | 0.18 (-0.06, 0.43) | 0.16 (-0.08, 0.4) | 0.16 (-0.08, 0.41) | 0.14 (-0.11, 0.38) | 0.16 (-0.09, 0.4) |
|  | Common Confounders | 0.07 (-0.26, 0.39) | 0.18 (-0.06, 0.42) | 0.16 (-0.08, 0.4) | 0.16 (-0.08, 0.4) | 0.14 (-0.1, 0.38) | 0.16 (-0.08, 0.4) |
| IPV Physical | Extended Confounder Set | 0.14 (-0.23, 0.5) | 0.28 (0.01, 0.55) | 0.21 (-0.06, 0.48) | 0.24 (-0.03, 0.5) | 0.16 (-0.11, 0.42) | 0.29 (0.02, 0.55) |
|  | Common Confounders | 0.13 (-0.23, 0.49) | 0.27 (0.01, 0.54) | 0.23 (-0.04, 0.49) | 0.23 (-0.03, 0.49) | 0.16 (-0.1, 0.42) | 0.29 (0.03, 0.55) |
| LEQ | Extended Confounder Set | 0.07 (-0.09, 0.24) | 0.05 (-0.07, 0.18) | 0.04 (-0.08, 0.16) | 0.03 (-0.09, 0.15) | 0.08 (-0.04, 0.19) | 0.07 (-0.05, 0.19) |
|  | Common Confounders | 0.07 (-0.1, 0.23) | 0.05 (-0.07, 0.17) | 0.04 (-0.08, 0.16) | 0.02 (-0.1, 0.14) | 0.07 (-0.04, 0.19) | 0.07 (-0.05, 0.19) |
| EPDS | Extended Confounder Set | 0.1 (-0.06, 0.26) | **0.17 (0.05, 0.29)** | **0.12 (0, 0.25)** | **0.14 (0.02, 0.26)** | **0.16 (0.04, 0.28)** | **0.16 (0.03, 0.28)** |
|  | Common Confounders | 0.11 (-0.06, 0.27) | **0.17 (0.05, 0.29)** | **0.13 (0.01, 0.25)** | **0.14 (0.02, 0.26)** | **0.16 (0.04, 0.28)** | **0.16 (0.04, 0.28)** |
| ASSIST Tobacco | Extended Confounder Set | 0.01 (-0.13, 0.14) | -0.03 (-0.11, 0.05) | -0.01 (-0.1, 0.08) | 0 (-0.09, 0.09) | 0.02 (-0.08, 0.11) | 0.02 (-0.08, 0.11) |
|  | Common Confounders | -0.06 (-0.15, 0.03) | -0.01 (-0.08, 0.05) | 0.01 (-0.06, 0.07) | 0.01 (-0.06, 0.07) | 0.01 (-0.05, 0.08) | 0.02 (-0.04, 0.09) |
| ASSIST Alcohol | Extended Confounder Set | **0.14 (0.03, 0.25)** | **0.12 (0.03, 0.2)** | **0.12 (0.04, 0.2)** | **0.11 (0.03, 0.19)** | **0.11 (0.03, 0.19)** | **0.11 (0.03, 0.19)** |
|  | Common Confounders | **0.11 (0, 0.22)** | **0.12 (0.04, 0.2)** | **0.12 (0.04, 0.2)** | **0.1 (0.03, 0.18)** | **0.11 (0.03, 0.19)** | **0.11 (0.03, 0.19)** |
| Inverse SES | Extended Confounder Set | 0.15 (-0.28, 0.58) | -0.01 (-0.32, 0.3) | -0.01 (-0.32, 0.3) | 0.01 (-0.31, 0.32) | 0.02 (-0.29, 0.33) | -0.02 (-0.33, 0.29) |
|  | Common Confounders | 0.21 (-0.21, 0.62) | 0.05 (-0.24, 0.35) | 0.05 (-0.24, 0.35) | 0.05 (-0.24, 0.35) | 0.05 (-0.24, 0.35) | 0.05 (-0.24, 0.35) |
|  |  | CBCL Externalizing Problems | | | | | |
| PM10 | Extended Confounder Set | 0.03 (-0.04, 0.1) | 0.02 (-0.03, 0.07) | 0.03 (-0.02, 0.08) | 0.03 (-0.02, 0.08) | 0.02 (-0.03, 0.07) | 0.04 (0, 0.09) |
|  | Common Confounders | 0.03 (-0.04, 0.1) | 0.03 (-0.02, 0.07) | 0.03 (-0.02, 0.08) | 0.03 (-0.02, 0.08) | 0.02 (-0.03, 0.07) | 0.04 (-0.01, 0.09) |
| CO | Extended Confounder Set | 0.03 (-0.01, 0.06) | 0.02 (-0.01, 0.04) | 0.01 (-0.02, 0.03) | 0.02 (-0.01, 0.04) | 0.02 (-0.01, 0.04) | 0.02 (-0.01, 0.05) |
|  | Common Confounders | 0.03 (-0.01, 0.06) | 0.02 (-0.01, 0.04) | 0.01 (-0.02, 0.03) | 0.02 (-0.01, 0.04) | 0.02 (-0.01, 0.04) | 0.02 (-0.01, 0.05) |
| Benzene | Extended Confounder Set | 0.05 (-0.02, 0.12) | 0.04 (-0.01, 0.1) | 0.04 (-0.01, 0.09) | **0.06 (0, 0.11)** | **0.06 (0.01, 0.11)** | 0.05 (0, 0.1) |
|  | Common Confounders | 0.05 (-0.02, 0.12) | 0.04 (-0.01, 0.1) | 0.04 (-0.01, 0.09) | **0.05 (0, 0.11)** | **0.06 (0, 0.11)** | 0.05 (0, 0.1) |
| Toluene | Extended Confounder Set | 0.05 (-0.01, 0.11) | 0.04 (-0.01, 0.09) | 0.04 (-0.01, 0.08) | **0.05 (0, 0.1)** | 0.04 (-0.01, 0.08) | 0.05 (0, 0.09) |
|  | Common Confounders | 0.05 (-0.01, 0.11) | 0.04 (-0.01, 0.09) | 0.04 (-0.01, 0.09) | **0.05 (0, 0.1)** | 0.04 (-0.01, 0.08) | **0.05 (0, 0.1)** |
| NO2 | Extended Confounder Set | 0.08 (-0.02, 0.17) | 0.05 (-0.02, 0.13) | 0.04 (-0.03, 0.11) | 0.06 (-0.01, 0.13) | 0.05 (-0.03, 0.12) | 0.05 (-0.03, 0.12) |
|  | Common Confounders | 0.06 (-0.03, 0.16) | 0.04 (-0.03, 0.12) | 0.03 (-0.04, 0.1) | 0.05 (-0.02, 0.12) | 0.04 (-0.03, 0.11) | 0.04 (-0.03, 0.11) |
| SO2 | Extended Confounder Set | 0.09 (-0.06, 0.25) | -0.02 (-0.12, 0.09) | -0.03 (-0.14, 0.08) | 0.01 (-0.1, 0.12) | -0.01 (-0.12, 0.1) | -0.01 (-0.12, 0.1) |
|  | Common Confounders | 0.08 (-0.07, 0.23) | -0.03 (-0.13, 0.07) | -0.05 (-0.15, 0.06) | 0 (-0.11, 0.11) | -0.03 (-0.14, 0.08) | -0.02 (-0.13, 0.09) |
| Cotinine | Extended Confounder Set | -0.1 (-0.19, -0.02) | -0.02 (-0.08, 0.04) | -0.02 (-0.08, 0.04) | -0.03 (-0.08, 0.03) | -0.03 (-0.09, 0.03) | -0.03 (-0.09, 0.03) |
|  | Common Confounders | -0.08 (-0.15, -0.01) | -0.01 (-0.06, 0.04) | 0 (-0.06, 0.05) | -0.01 (-0.06, 0.04) | -0.01 (-0.06, 0.05) | 0 (-0.06, 0.05) |
| Food Insecurity | Extended Confounder Set | -0.03 (-0.2, 0.15) | 0.04 (-0.08, 0.17) | 0.03 (-0.1, 0.15) | 0 (-0.13, 0.12) | 0.04 (-0.08, 0.17) | 0.02 (-0.11, 0.14) |
|  | Common Confounders | -0.05 (-0.22, 0.13) | 0.05 (-0.08, 0.17) | 0.03 (-0.09, 0.15) | 0 (-0.12, 0.13) | 0.04 (-0.08, 0.17) | 0.02 (-0.11, 0.14) |
| SRQ | Extended Confounder Set | 0.11 (-0.02, 0.24) | **0.1 (0.01, 0.19)** | **0.09 (0, 0.19)** | 0.1 (0, 0.19) | 0.09 (0, 0.18) | 0.08 (-0.02, 0.17) |
|  | Common Confounders | 0.1 (-0.03, 0.23) | 0.09 (0, 0.18) | 0.09 (-0.01, 0.18) | 0.09 (0, 0.18) | 0.08 (-0.01, 0.17) | 0.07 (-0.02, 0.16) |
| IPV Emotional | Extended Confounder Set | 0.12 (-0.18, 0.43) | 0.16 (-0.07, 0.38) | 0.14 (-0.08, 0.37) | 0.12 (-0.1, 0.35) | 0.12 (-0.11, 0.34) | 0.15 (-0.08, 0.37) |
|  | Common Confounders | 0.07 (-0.23, 0.37) | 0.15 (-0.08, 0.37) | 0.14 (-0.08, 0.36) | 0.11 (-0.11, 0.33) | 0.1 (-0.12, 0.32) | 0.14 (-0.08, 0.37) |
| IPV Physical | Extended Confounder Set | 0.14 (-0.2, 0.48) | 0.23 (-0.02, 0.47) | 0.17 (-0.08, 0.42) | 0.19 (-0.06, 0.43) | 0.12 (-0.13, 0.36) | **0.24 (0, 0.49)** |
|  | Common Confounders | 0.11 (-0.22, 0.45) | 0.21 (-0.03, 0.45) | 0.17 (-0.07, 0.41) | 0.16 (-0.08, 0.4) | 0.1 (-0.14, 0.33) | 0.24 (0, 0.48) |
| LEQ | Extended Confounder Set | 0.08 (-0.08, 0.23) | 0.02 (-0.09, 0.13) | -0.01 (-0.12, 0.11) | 0.02 (-0.09, 0.13) | 0.07 (-0.04, 0.17) | 0.04 (-0.07, 0.16) |
|  | Common Confounders | 0.07 (-0.08, 0.23) | 0.01 (-0.1, 0.13) | -0.01 (-0.12, 0.1) | 0.01 (-0.1, 0.12) | 0.06 (-0.05, 0.17) | 0.04 (-0.07, 0.15) |
| EPDS | Extended Confounder Set | 0.05 (-0.1, 0.2) | **0.12 (0.01, 0.24)** | 0.09 (-0.03, 0.2) | 0.1 (-0.02, 0.21) | **0.12 (0.01, 0.23)** | 0.11 (0, 0.22) |
|  | Common Confounders | 0.05 (-0.1, 0.21) | **0.12 (0.01, 0.23)** | 0.09 (-0.02, 0.2) | 0.09 (-0.02, 0.2) | **0.11 (0, 0.22)** | 0.11 (0, 0.22) |
| ASSIST Tobacco | Extended Confounder Set | -0.01 (-0.14, 0.11) | -0.03 (-0.11, 0.04) | -0.02 (-0.11, 0.06) | -0.01 (-0.1, 0.07) | 0.01 (-0.08, 0.1) | 0.01 (-0.08, 0.1) |
|  | Common Confounders | -0.07 (-0.16, 0.01) | -0.02 (-0.08, 0.04) | -0.01 (-0.07, 0.05) | -0.01 (-0.07, 0.05) | 0.01 (-0.06, 0.07) | 0.01 (-0.05, 0.07) |
| ASSIST Alcohol | Extended Confounder Set | **0.14 (0.04, 0.24)** | **0.11 (0.03, 0.19)** | **0.11 (0.03, 0.18)** | **0.1 (0.03, 0.18)** | **0.11 (0.03, 0.18)** | **0.11 (0.04, 0.19)** |
|  | Common Confounders | **0.11 (0.01, 0.2)** | **0.11 (0.03, 0.18)** | **0.1 (0.03, 0.18)** | **0.1 (0.03, 0.17)** | **0.1 (0.03, 0.18)** | **0.11 (0.04, 0.18)** |
| Inverse SES | Extended Confounder Set | 0.08 (-0.32, 0.48) | -0.01 (-0.29, 0.28) | -0.01 (-0.3, 0.27) | 0 (-0.29, 0.29) | 0.03 (-0.25, 0.32) | -0.02 (-0.31, 0.26) |
|  | Common Confounders | 0.1 (-0.28, 0.48) | 0.02 (-0.25, 0.3) | 0.02 (-0.25, 0.3) | 0.02 (-0.25, 0.3) | 0.02 (-0.25, 0.3) | 0.02 (-0.25, 0.3) |
|  |  | CBCL Internalizing Problems | | | | | |
| PM10 | Extended Confounder Set | 0.05 (-0.01, 0.1) | 0.04 (0, 0.08) | 0.04 (0, 0.08) | 0.04 (0, 0.08) | 0.03 (-0.01, 0.07) | **0.04 (0, 0.08)** |
|  | Common Confounders | 0.05 (-0.01, 0.1) | **0.04 (0, 0.08)** | 0.04 (0, 0.08) | **0.04 (0, 0.08)** | 0.03 (-0.01, 0.07) | **0.04 (0, 0.08)** |
| CO | Extended Confounder Set | 0 (-0.02, 0.03) | -0.01 (-0.03, 0.01) | 0 (-0.02, 0.02) | -0.01 (-0.03, 0.01) | -0.01 (-0.03, 0.01) | 0 (-0.03, 0.02) |
|  | Common Confounders | 0 (-0.02, 0.03) | -0.01 (-0.03, 0.01) | 0 (-0.02, 0.02) | -0.01 (-0.03, 0.02) | 0 (-0.03, 0.02) | 0 (-0.02, 0.02) |
| Benzene | Extended Confounder Set | -0.01 (-0.07, 0.05) | -0.02 (-0.06, 0.03) | -0.02 (-0.06, 0.03) | -0.01 (-0.05, 0.03) | -0.02 (-0.06, 0.03) | -0.01 (-0.05, 0.04) |
|  | Common Confounders | -0.01 (-0.06, 0.05) | -0.01 (-0.06, 0.03) | -0.01 (-0.06, 0.03) | -0.01 (-0.05, 0.03) | -0.02 (-0.06, 0.03) | 0 (-0.05, 0.04) |
| Toluene | Extended Confounder Set | -0.02 (-0.07, 0.03) | -0.01 (-0.05, 0.02) | -0.02 (-0.06, 0.02) | -0.01 (-0.05, 0.02) | -0.02 (-0.06, 0.02) | 0 (-0.04, 0.03) |
|  | Common Confounders | -0.02 (-0.07, 0.03) | -0.01 (-0.05, 0.03) | -0.01 (-0.05, 0.02) | -0.01 (-0.05, 0.03) | -0.02 (-0.06, 0.02) | 0 (-0.04, 0.04) |
| NO2 | Extended Confounder Set | 0.06 (-0.01, 0.14) | 0.04 (-0.02, 0.1) | 0.01 (-0.05, 0.06) | 0.02 (-0.03, 0.08) | 0.04 (-0.02, 0.1) | 0.03 (-0.03, 0.09) |
|  | Common Confounders | 0.06 (-0.02, 0.13) | 0.03 (-0.03, 0.09) | 0 (-0.05, 0.06) | 0.02 (-0.04, 0.07) | 0.03 (-0.02, 0.09) | 0.03 (-0.03, 0.08) |
| SO2 | Extended Confounder Set | 0.11 (-0.01, 0.23) | -0.02 (-0.1, 0.06) | -0.01 (-0.1, 0.08) | 0 (-0.09, 0.09) | -0.01 (-0.1, 0.08) | -0.01 (-0.1, 0.07) |
|  | Common Confounders | 0.09 (-0.04, 0.21) | -0.04 (-0.12, 0.04) | -0.03 (-0.11, 0.06) | -0.02 (-0.11, 0.07) | -0.02 (-0.11, 0.06) | -0.03 (-0.12, 0.06) |
| Cotinine | Extended Confounder Set | -0.02 (-0.09, 0.05) | 0 (-0.05, 0.05) | -0.01 (-0.06, 0.04) | 0 (-0.04, 0.05) | 0 (-0.05, 0.05) | 0 (-0.05, 0.05) |
|  | Common Confounders | -0.01 (-0.07, 0.05) | 0.01 (-0.04, 0.05) | 0.01 (-0.04, 0.05) | 0.01 (-0.03, 0.05) | 0 (-0.04, 0.04) | 0.01 (-0.04, 0.05) |
| Food Insecurity | Extended Confounder Set | -0.01 (-0.15, 0.13) | 0.07 (-0.03, 0.17) | 0.05 (-0.05, 0.15) | 0.05 (-0.05, 0.15) | 0.03 (-0.06, 0.13) | 0.07 (-0.03, 0.17) |
|  | Common Confounders | -0.02 (-0.16, 0.12) | 0.07 (-0.03, 0.17) | 0.05 (-0.05, 0.15) | 0.05 (-0.05, 0.16) | 0.03 (-0.06, 0.13) | 0.07 (-0.03, 0.17) |
| SRQ | Extended Confounder Set | **0.13 (0.03, 0.23)** | **0.11 (0.03, 0.18)** | **0.08 (0.01, 0.16)** | **0.1 (0.03, 0.18)** | **0.1 (0.03, 0.18)** | **0.1 (0.02, 0.17)** |
|  | Common Confounders | **0.11 (0.01, 0.21)** | **0.1 (0.03, 0.17)** | **0.08 (0, 0.15)** | **0.1 (0.02, 0.17)** | **0.1 (0.03, 0.17)** | **0.09 (0.02, 0.17)** |
| IPV Emotional | Extended Confounder Set | -0.08 (-0.32, 0.17) | -0.01 (-0.19, 0.17) | -0.01 (-0.19, 0.17) | -0.01 (-0.19, 0.17) | 0.01 (-0.17, 0.19) | 0 (-0.18, 0.19) |
|  | Common Confounders | -0.08 (-0.32, 0.16) | 0 (-0.18, 0.18) | 0 (-0.18, 0.18) | 0 (-0.18, 0.18) | 0.01 (-0.17, 0.19) | 0.01 (-0.17, 0.19) |
| IPV Physical | Extended Confounder Set | 0.08 (-0.19, 0.35) | 0.09 (-0.11, 0.29) | 0.09 (-0.11, 0.29) | 0.12 (-0.07, 0.32) | 0.07 (-0.13, 0.26) | 0.15 (-0.05, 0.34) |
|  | Common Confounders | 0.11 (-0.16, 0.37) | 0.1 (-0.09, 0.3) | 0.11 (-0.09, 0.3) | 0.13 (-0.06, 0.32) | 0.07 (-0.12, 0.26) | 0.15 (-0.04, 0.35) |
| LEQ | Extended Confounder Set | 0.01 (-0.11, 0.14) | 0 (-0.09, 0.09) | 0.01 (-0.08, 0.1) | -0.02 (-0.11, 0.07) | -0.02 (-0.11, 0.07) | 0 (-0.09, 0.09) |
|  | Common Confounders | 0.02 (-0.11, 0.14) | 0 (-0.09, 0.09) | 0.01 (-0.08, 0.1) | -0.02 (-0.11, 0.07) | -0.01 (-0.1, 0.07) | 0 (-0.09, 0.09) |
| EPDS | Extended Confounder Set | 0.04 (-0.08, 0.16) | 0.05 (-0.04, 0.14) | 0.04 (-0.05, 0.13) | 0.05 (-0.04, 0.15) | 0.07 (-0.02, 0.16) | 0.06 (-0.03, 0.15) |
|  | Common Confounders | 0.05 (-0.07, 0.17) | 0.05 (-0.04, 0.14) | 0.04 (-0.05, 0.14) | 0.06 (-0.03, 0.15) | 0.07 (-0.02, 0.16) | 0.06 (-0.03, 0.15) |
| ASSIST Tobacco | Extended Confounder Set | 0 (-0.1, 0.1) | -0.01 (-0.07, 0.05) | -0.01 (-0.08, 0.06) | -0.02 (-0.09, 0.05) | -0.02 (-0.09, 0.05) | -0.02 (-0.09, 0.05) |
|  | Common Confounders | 0 (-0.07, 0.06) | 0 (-0.05, 0.05) | 0 (-0.05, 0.05) | 0 (-0.05, 0.05) | 0 (-0.05, 0.05) | 0 (-0.05, 0.05) |
| ASSIST Alcohol | Extended Confounder Set | **0.1 (0.02, 0.18)** | **0.07 (0.01, 0.13)** | **0.07 (0.01, 0.13)** | **0.07 (0.01, 0.13)** | **0.07 (0.01, 0.14)** | **0.07 (0.01, 0.13)** |
|  | Common Confounders | **0.09 (0.01, 0.17)** | **0.07 (0.01, 0.13)** | **0.07 (0.01, 0.13)** | **0.07 (0.02, 0.13)** | **0.07 (0.01, 0.13)** | **0.07 (0.01, 0.13)** |
| Inverse SES | Extended Confounder Set | 0.02 (-0.3, 0.34) | 0.01 (-0.22, 0.24) | 0.02 (-0.22, 0.25) | 0.02 (-0.21, 0.26) | 0.04 (-0.19, 0.27) | 0.03 (-0.2, 0.26) |
|  | Common Confounders | 0.09 (-0.21, 0.4) | 0.06 (-0.16, 0.28) | 0.06 (-0.16, 0.28) | 0.06 (-0.16, 0.28) | 0.06 (-0.16, 0.28) | 0.06 (-0.16, 0.28) |

**Table S4**. Beta estimates and 95% CIs for individual postnatal exposure adjusted linear regression models. The common confounders linear regression models were adjusted for maternal HIV status, maternal age, ancestry, and socioeconomic status. Extended confounder set models using indoor air pollutant exposures were additionally adjusted principal components of psychosocial factors, and vice versa. Tables shows results from complete case models as well as multiple imputation (MI) models using 5 different random seeds (MI1 to MI5). MI5 models were presented in the main analysis.

|  |  | Complete Case | MI1 | MI2 | MI3 | MI4 | MI5 (Analysis Sample) |
| --- | --- | --- | --- | --- | --- | --- | --- |
|  |  | CBCL Total Problems | | | | | |
| PM10 | Extended Confounder Set | 0.05 (-0.09, 0.18) | -0.02 (-0.07, 0.03) | -0.01 (-0.06, 0.04) | -0.01 (-0.06, 0.05) | -0.03 (-0.08, 0.02) | -0.03 (-0.09, 0.02) |
|  | Common Confounders | 0.04 (-0.1, 0.17) | -0.02 (-0.07, 0.03) | -0.01 (-0.06, 0.04) | 0 (-0.06, 0.05) | -0.03 (-0.08, 0.02) | -0.03 (-0.09, 0.02) |
| CO | Extended Confounder Set | 0.06 (-0.01, 0.14) | 0.03 (0, 0.05) | **0.03 (0, 0.06)** | **0.04 (0.01, 0.07)** | 0.02 (-0.01, 0.05) | **0.03 (0, 0.06)** |
|  | Common Confounders | 0.06 (-0.02, 0.13) | 0.03 (0, 0.05) | **0.03 (0, 0.06)** | **0.03 (0.01, 0.06)** | 0.02 (-0.01, 0.05) | **0.03 (0, 0.06)** |
| Benzene | Extended Confounder Set | -0.07 (-0.24, 0.1) | 0 (-0.06, 0.05) | -0.02 (-0.07, 0.04) | -0.05 (-0.11, 0.01) | -0.03 (-0.1, 0.03) | -0.01 (-0.07, 0.05) |
|  | Common Confounders | -0.06 (-0.23, 0.1) | 0 (-0.06, 0.05) | -0.01 (-0.07, 0.04) | -0.04 (-0.1, 0.02) | -0.03 (-0.09, 0.03) | -0.01 (-0.06, 0.05) |
| Toluene | Extended Confounder Set | -0.04 (-0.17, 0.09) | 0 (-0.04, 0.05) | 0 (-0.05, 0.05) | -0.02 (-0.07, 0.03) | -0.02 (-0.07, 0.03) | -0.02 (-0.07, 0.03) |
|  | Common Confounders | -0.05 (-0.18, 0.08) | 0 (-0.05, 0.05) | 0 (-0.05, 0.05) | -0.01 (-0.06, 0.04) | -0.02 (-0.07, 0.03) | -0.02 (-0.07, 0.03) |
| NO2 | Extended Confounder Set | -0.01 (-0.25, 0.23) | 0 (-0.09, 0.09) | 0.01 (-0.08, 0.09) | 0.04 (-0.04, 0.13) | -0.02 (-0.1, 0.06) | 0.04 (-0.04, 0.12) |
|  | Common Confounders | 0 (-0.23, 0.23) | 0 (-0.09, 0.09) | 0.01 (-0.08, 0.09) | 0.05 (-0.03, 0.13) | -0.01 (-0.09, 0.06) | 0.04 (-0.04, 0.12) |
| SO2 | Extended Confounder Set | -0.11 (-0.56, 0.34) | -0.05 (-0.19, 0.09) | -0.06 (-0.2, 0.08) | -0.11 (-0.22, -0.01) | -0.08 (-0.21, 0.05) | -0.08 (-0.2, 0.04) |
|  | Common Confounders | -0.06 (-0.49, 0.37) | -0.05 (-0.18, 0.09) | -0.07 (-0.21, 0.07) | -0.12 (-0.22, -0.02) | -0.08 (-0.21, 0.04) | -0.08 (-0.19, 0.04) |
| Food Insecurity | Extended Confounder Set | 0.01 (-0.57, 0.59) | -0.06 (-0.26, 0.14) | -0.02 (-0.22, 0.18) | 0.04 (-0.15, 0.23) | 0.04 (-0.15, 0.23) | -0.02 (-0.22, 0.17) |
|  | Common Confounders | -0.05 (-0.57, 0.47) | -0.06 (-0.25, 0.14) | -0.03 (-0.23, 0.18) | 0.03 (-0.16, 0.22) | 0.04 (-0.14, 0.23) | -0.02 (-0.21, 0.17) |
| SRQ | Extended Confounder Set | 0.2 (-0.08, 0.48) | 0.04 (-0.06, 0.14) | **0.1 (0, 0.19)** | 0.04 (-0.06, 0.14) | 0.05 (-0.04, 0.15) | 0 (-0.09, 0.1) |
|  | Common Confounders | 0.19 (-0.08, 0.46) | 0.04 (-0.06, 0.14) | **0.1 (0.01, 0.2)** | 0.03 (-0.06, 0.13) | 0.05 (-0.04, 0.15) | 0 (-0.09, 0.1) |
| IPV Emotional | Extended Confounder Set | 0 (-0.76, 0.76) | -0.03 (-0.27, 0.22) | 0.17 (-0.08, 0.43) | 0.08 (-0.14, 0.29) | 0.06 (-0.19, 0.31) | 0.03 (-0.23, 0.28) |
|  | Common Confounders | 0.01 (-0.73, 0.74) | -0.03 (-0.27, 0.21) | 0.18 (-0.07, 0.44) | 0.07 (-0.15, 0.29) | 0.05 (-0.2, 0.29) | 0.03 (-0.22, 0.28) |
| IPV Physical | Extended Confounder Set | 0.15 (-0.76, 1.07) | -0.01 (-0.29, 0.27) | 0.09 (-0.21, 0.39) | 0.09 (-0.17, 0.35) | 0.17 (-0.1, 0.44) | 0 (-0.28, 0.28) |
|  | Common Confounders | 0.2 (-0.67, 1.07) | -0.01 (-0.29, 0.27) | 0.1 (-0.2, 0.39) | 0.08 (-0.18, 0.34) | 0.15 (-0.12, 0.42) | 0 (-0.27, 0.28) |
| LEQ | Extended Confounder Set | 0.02 (-0.39, 0.43) | 0.02 (-0.1, 0.15) | **0.14 (0.01, 0.27)** | 0.11 (-0.02, 0.24) | 0.08 (-0.05, 0.21) | -0.04 (-0.16, 0.08) |
|  | Common Confounders | 0.02 (-0.37, 0.41) | 0.02 (-0.1, 0.15) | **0.14 (0.02, 0.27)** | 0.11 (-0.02, 0.23) | 0.08 (-0.05, 0.21) | -0.04 (-0.16, 0.08) |
| EPDS | Extended Confounder Set | 0.27 (-0.11, 0.64) | 0.08 (-0.03, 0.19) | **0.15 (0.04, 0.26)** | **0.14 (0.03, 0.24)** | 0.09 (-0.02, 0.2) | 0.1 (0, 0.21) |
|  | Common Confounders | 0.28 (-0.09, 0.64) | 0.08 (-0.03, 0.19) | **0.15 (0.05, 0.26)** | **0.14 (0.03, 0.24)** | 0.09 (-0.01, 0.19) | 0.1 (0, 0.2) |
| ASSIST Tobacco | Extended Confounder Set | -0.16 (-0.38, 0.06) | 0 (-0.06, 0.07) | 0.01 (-0.06, 0.08) | 0.03 (-0.07, 0.12) | 0 (-0.07, 0.08) | -0.02 (-0.07, 0.03) |
|  | Common Confounders | -0.16 (-0.38, 0.05) | 0 (-0.06, 0.07) | 0.01 (-0.06, 0.08) | 0.03 (-0.06, 0.12) | 0 (-0.07, 0.08) | -0.02 (-0.08, 0.03) |
| ASSIST Alcohol | Extended Confounder Set | -0.16 (-0.38, 0.06) | 0.01 (-0.06, 0.08) | 0 (-0.06, 0.06) | 0.03 (-0.07, 0.12) | 0.01 (-0.08, 0.1) | 0.05 (-0.02, 0.13) |
|  | Common Confounders | -0.16 (-0.38, 0.05) | 0.01 (-0.06, 0.08) | 0 (-0.06, 0.06) | 0.03 (-0.06, 0.12) | 0.01 (-0.08, 0.1) | 0.05 (-0.03, 0.13) |
| Inverse SES | Extended Confounder Set | 0.1 (-0.71, 0.9) | -0.03 (-0.3, 0.24) | 0.08 (-0.19, 0.34) | -0.06 (-0.33, 0.21) | -0.12 (-0.37, 0.13) | 0.01 (-0.25, 0.27) |
|  | Common Confounders | 0.11 (-0.67, 0.89) | -0.03 (-0.29, 0.24) | 0.08 (-0.19, 0.34) | -0.06 (-0.32, 0.21) | -0.11 (-0.36, 0.14) | 0.01 (-0.25, 0.27) |
|  |  | CBCL Externalizing Problems | | | | | |
| PM10 | Extended Confounder Set | 0.02 (-0.1, 0.13) | -0.01 (-0.06, 0.04) | 0 (-0.05, 0.05) | 0 (-0.05, 0.05) | -0.01 (-0.05, 0.04) | -0.01 (-0.06, 0.04) |
|  | Common Confounders | 0 (-0.11, 0.12) | -0.01 (-0.06, 0.04) | 0 (-0.05, 0.05) | 0 (-0.05, 0.05) | -0.01 (-0.05, 0.04) | -0.01 (-0.06, 0.03) |
| CO | Extended Confounder Set | 0.06 (-0.01, 0.12) | **0.03 (0.01, 0.06)** | **0.03 (0.01, 0.06)** | **0.03 (0, 0.06)** | 0.02 (-0.01, 0.04) | **0.03 (0, 0.06)** |
|  | Common Confounders | 0.05 (-0.01, 0.12) | **0.03 (0.01, 0.05)** | **0.03 (0.01, 0.06)** | **0.03 (0, 0.06)** | 0.02 (-0.01, 0.04) | **0.03 (0, 0.06)** |
| Benzene | Extended Confounder Set | -0.03 (-0.17, 0.12) | 0.01 (-0.04, 0.06) | -0.01 (-0.06, 0.04) | -0.03 (-0.08, 0.03) | -0.01 (-0.07, 0.04) | 0.01 (-0.04, 0.07) |
|  | Common Confounders | -0.01 (-0.15, 0.14) | 0.01 (-0.04, 0.06) | -0.01 (-0.06, 0.05) | -0.02 (-0.08, 0.03) | -0.01 (-0.06, 0.05) | 0.02 (-0.04, 0.07) |
| Toluene | Extended Confounder Set | -0.01 (-0.12, 0.11) | 0.02 (-0.02, 0.06) | 0.01 (-0.03, 0.06) | -0.01 (-0.06, 0.04) | -0.01 (-0.06, 0.04) | -0.01 (-0.05, 0.04) |
|  | Common Confounders | 0 (-0.11, 0.11) | 0.02 (-0.03, 0.06) | 0.01 (-0.03, 0.06) | 0 (-0.05, 0.05) | -0.01 (-0.05, 0.04) | -0.01 (-0.05, 0.04) |
| NO2 | Extended Confounder Set | 0.02 (-0.19, 0.22) | 0.02 (-0.06, 0.1) | 0.02 (-0.06, 0.09) | 0.07 (0, 0.15) | -0.01 (-0.08, 0.06) | 0.05 (-0.02, 0.13) |
|  | Common Confounders | 0.05 (-0.14, 0.25) | 0.02 (-0.06, 0.1) | 0.01 (-0.06, 0.09) | 0.08 (0, 0.15) | -0.01 (-0.08, 0.06) | 0.05 (-0.02, 0.13) |
| SO2 | Extended Confounder Set | -0.29 (-0.67, 0.08) | -0.1 (-0.23, 0.03) | -0.13 (-0.27, 0) | -0.09 (-0.18, 0) | -0.1 (-0.22, 0.02) | -0.09 (-0.2, 0.01) |
|  | Common Confounders | -0.31 (-0.67, 0.06) | -0.1 (-0.22, 0.03) | -0.14 (-0.27, -0.01) | -0.09 (-0.19, 0) | -0.1 (-0.22, 0.01) | -0.1 (-0.2, 0.01) |
| Food Insecurity | Extended Confounder Set | -0.27 (-0.75, 0.21) | -0.04 (-0.22, 0.14) | 0.02 (-0.16, 0.21) | 0.02 (-0.16, 0.19) | 0.05 (-0.12, 0.22) | 0 (-0.18, 0.17) |
|  | Common Confounders | -0.41 (-0.85, 0.02) | -0.04 (-0.23, 0.14) | 0.01 (-0.17, 0.2) | 0.02 (-0.16, 0.19) | 0.05 (-0.12, 0.22) | 0 (-0.17, 0.18) |
| SRQ | Extended Confounder Set | 0.18 (-0.05, 0.41) | 0.04 (-0.05, 0.13) | 0.08 (-0.01, 0.17) | 0.04 (-0.05, 0.13) | 0.04 (-0.05, 0.13) | 0.02 (-0.07, 0.11) |
|  | Common Confounders | 0.16 (-0.07, 0.39) | 0.04 (-0.05, 0.13) | 0.09 (0, 0.17) | 0.04 (-0.05, 0.12) | 0.04 (-0.05, 0.13) | 0.03 (-0.06, 0.11) |
| IPV Emotional | Extended Confounder Set | -0.01 (-0.65, 0.62) | -0.02 (-0.24, 0.2) | 0.1 (-0.13, 0.34) | 0.07 (-0.13, 0.27) | 0.02 (-0.2, 0.25) | 0.03 (-0.2, 0.26) |
|  | Common Confounders | 0.01 (-0.62, 0.64) | -0.02 (-0.24, 0.2) | 0.12 (-0.11, 0.35) | 0.06 (-0.14, 0.26) | 0.02 (-0.21, 0.25) | 0.05 (-0.18, 0.28) |
| IPV Physical | Extended Confounder Set | -0.17 (-0.93, 0.59) | -0.06 (-0.31, 0.2) | 0.06 (-0.21, 0.34) | 0.05 (-0.19, 0.28) | 0.1 (-0.15, 0.35) | 0 (-0.25, 0.26) |
|  | Common Confounders | -0.07 (-0.82, 0.67) | -0.06 (-0.31, 0.2) | 0.08 (-0.19, 0.35) | 0.04 (-0.2, 0.28) | 0.08 (-0.17, 0.33) | 0.01 (-0.24, 0.27) |
| LEQ | Extended Confounder Set | -0.04 (-0.38, 0.29) | -0.03 (-0.14, 0.09) | 0.08 (-0.03, 0.2) | 0.05 (-0.07, 0.17) | 0.05 (-0.07, 0.17) | -0.04 (-0.15, 0.07) |
|  | Common Confounders | -0.06 (-0.39, 0.28) | -0.03 (-0.14, 0.09) | 0.09 (-0.03, 0.2) | 0.04 (-0.07, 0.16) | 0.04 (-0.08, 0.16) | -0.04 (-0.15, 0.07) |
| EPDS | Extended Confounder Set | 0.18 (-0.13, 0.5) | **0.11 (0.01, 0.21)** | **0.14 (0.04, 0.24)** | **0.14 (0.04, 0.23)** | **0.1 (0, 0.2)** | **0.13 (0.03, 0.23)** |
|  | Common Confounders | 0.2 (-0.11, 0.52) | **0.11 (0.01, 0.21)** | **0.14 (0.05, 0.24)** | **0.14 (0.04, 0.23)** | **0.1 (0.01, 0.2)** | **0.13 (0.03, 0.22)** |
| ASSIST Tobacco | Extended Confounder Set | -0.08 (-0.26, 0.11) | 0.01 (-0.05, 0.07) | 0.01 (-0.05, 0.08) | 0.03 (-0.05, 0.12) | 0.02 (-0.05, 0.08) | -0.02 (-0.07, 0.03) |
|  | Common Confounders | -0.08 (-0.26, 0.11) | 0.01 (-0.05, 0.07) | 0.02 (-0.05, 0.08) | 0.04 (-0.05, 0.12) | 0.02 (-0.05, 0.09) | -0.02 (-0.07, 0.03) |
| ASSIST Alcohol | Extended Confounder Set | -0.08 (-0.26, 0.11) | 0.01 (-0.05, 0.08) | 0 (-0.05, 0.06) | 0.03 (-0.05, 0.12) | 0.02 (-0.06, 0.11) | 0.05 (-0.02, 0.13) |
|  | Common Confounders | -0.08 (-0.26, 0.11) | 0.01 (-0.05, 0.08) | 0.01 (-0.05, 0.06) | 0.04 (-0.05, 0.12) | 0.03 (-0.06, 0.11) | 0.05 (-0.02, 0.12) |
| Inverse SES | Extended Confounder Set | -0.53 (-1.19, 0.13) | -0.03 (-0.28, 0.21) | 0.01 (-0.24, 0.25) | -0.13 (-0.38, 0.11) | -0.14 (-0.37, 0.09) | -0.05 (-0.28, 0.19) |
|  | Common Confounders | -0.49 (-1.15, 0.17) | -0.04 (-0.28, 0.2) | 0 (-0.24, 0.25) | -0.13 (-0.38, 0.12) | -0.13 (-0.36, 0.1) | -0.04 (-0.28, 0.2) |
|  |  | CBCL Internalizing Problems | | | | | |
| PM10 | Extended Confounder Set | 0.09 (-0.03, 0.2) | 0.01 (-0.03, 0.04) | 0.03 (-0.01, 0.07) | 0.01 (-0.03, 0.05) | 0.01 (-0.03, 0.05) | -0.02 (-0.05, 0.02) |
|  | Common Confounders | 0.09 (-0.02, 0.19) | 0.01 (-0.03, 0.04) | 0.03 (-0.01, 0.07) | 0.01 (-0.03, 0.05) | 0.01 (-0.03, 0.05) | -0.01 (-0.05, 0.03) |
| CO | Extended Confounder Set | 0.04 (-0.02, 0.11) | 0.01 (-0.01, 0.03) | 0 (-0.02, 0.02) | 0.01 (-0.01, 0.03) | 0 (-0.02, 0.02) | 0.01 (-0.01, 0.03) |
|  | Common Confounders | 0.04 (-0.02, 0.1) | 0.01 (-0.01, 0.03) | 0 (-0.02, 0.02) | 0.01 (-0.01, 0.03) | 0 (-0.02, 0.02) | 0.01 (-0.01, 0.03) |
| Benzene | Extended Confounder Set | 0 (-0.14, 0.14) | -0.02 (-0.06, 0.03) | -0.01 (-0.05, 0.03) | -0.03 (-0.07, 0.02) | -0.02 (-0.07, 0.02) | 0.01 (-0.04, 0.05) |
|  | Common Confounders | -0.01 (-0.15, 0.13) | -0.02 (-0.06, 0.03) | -0.01 (-0.05, 0.04) | -0.02 (-0.07, 0.02) | -0.02 (-0.07, 0.02) | 0.01 (-0.03, 0.05) |
| Toluene | Extended Confounder Set | 0.03 (-0.08, 0.14) | -0.02 (-0.05, 0.02) | 0 (-0.04, 0.04) | -0.01 (-0.05, 0.03) | 0 (-0.04, 0.04) | 0 (-0.03, 0.04) |
|  | Common Confounders | 0.02 (-0.09, 0.12) | -0.02 (-0.05, 0.02) | 0 (-0.04, 0.04) | -0.01 (-0.04, 0.03) | 0 (-0.04, 0.04) | 0.01 (-0.03, 0.04) |
| NO2 | Extended Confounder Set | 0.09 (-0.11, 0.29) | 0 (-0.06, 0.07) | 0 (-0.07, 0.06) | 0.01 (-0.05, 0.08) | -0.02 (-0.07, 0.04) | 0.01 (-0.05, 0.07) |
|  | Common Confounders | 0.05 (-0.14, 0.24) | 0 (-0.06, 0.07) | 0 (-0.06, 0.06) | 0.02 (-0.04, 0.08) | -0.01 (-0.07, 0.04) | 0.01 (-0.05, 0.06) |
| SO2 | Extended Confounder Set | -0.14 (-0.51, 0.23) | -0.06 (-0.16, 0.04) | -0.01 (-0.11, 0.1) | -0.06 (-0.14, 0.01) | -0.04 (-0.14, 0.05) | -0.05 (-0.14, 0.03) |
|  | Common Confounders | -0.08 (-0.44, 0.28) | -0.06 (-0.16, 0.04) | -0.01 (-0.12, 0.1) | -0.06 (-0.14, 0.01) | -0.04 (-0.13, 0.05) | -0.06 (-0.14, 0.03) |
| Food Insecurity | Extended Confounder Set | 0.27 (-0.21, 0.74) | -0.07 (-0.21, 0.08) | -0.08 (-0.23, 0.07) | 0.04 (-0.11, 0.18) | 0 (-0.14, 0.13) | -0.03 (-0.17, 0.11) |
|  | Common Confounders | 0.16 (-0.27, 0.59) | -0.08 (-0.23, 0.06) | -0.08 (-0.23, 0.06) | 0.02 (-0.12, 0.16) | -0.01 (-0.14, 0.13) | -0.03 (-0.17, 0.11) |
| SRQ | Extended Confounder Set | 0.03 (-0.2, 0.26) | 0.04 (-0.03, 0.11) | 0.04 (-0.03, 0.12) | 0.02 (-0.05, 0.1) | -0.01 (-0.08, 0.07) | 0 (-0.07, 0.07) |
|  | Common Confounders | 0.03 (-0.19, 0.26) | 0.03 (-0.04, 0.1) | 0.05 (-0.02, 0.12) | 0.02 (-0.05, 0.09) | -0.01 (-0.08, 0.07) | 0 (-0.07, 0.07) |
| IPV Emotional | Extended Confounder Set | -0.12 (-0.74, 0.51) | -0.03 (-0.21, 0.15) | 0.06 (-0.13, 0.25) | 0.01 (-0.15, 0.17) | -0.03 (-0.21, 0.16) | -0.01 (-0.2, 0.18) |
|  | Common Confounders | -0.08 (-0.68, 0.53) | -0.02 (-0.2, 0.15) | 0.07 (-0.12, 0.25) | 0.02 (-0.14, 0.18) | -0.02 (-0.2, 0.17) | -0.01 (-0.19, 0.18) |
| IPV Physical | Extended Confounder Set | 0.21 (-0.54, 0.96) | 0.06 (-0.15, 0.27) | 0.11 (-0.11, 0.33) | 0.07 (-0.13, 0.26) | 0.12 (-0.08, 0.32) | 0.03 (-0.18, 0.23) |
|  | Common Confounders | 0.24 (-0.48, 0.96) | 0.05 (-0.16, 0.26) | 0.1 (-0.12, 0.32) | 0.07 (-0.12, 0.26) | 0.13 (-0.07, 0.33) | 0.03 (-0.18, 0.23) |
| LEQ | Extended Confounder Set | 0.04 (-0.3, 0.37) | 0.04 (-0.05, 0.14) | 0.07 (-0.02, 0.17) | 0.07 (-0.03, 0.16) | 0.02 (-0.07, 0.12) | -0.03 (-0.12, 0.06) |
|  | Common Confounders | 0.01 (-0.31, 0.33) | 0.04 (-0.06, 0.13) | 0.07 (-0.02, 0.17) | 0.06 (-0.04, 0.15) | 0.02 (-0.08, 0.11) | -0.03 (-0.12, 0.06) |
| EPDS | Extended Confounder Set | 0.11 (-0.2, 0.43) | -0.01 (-0.09, 0.07) | 0.05 (-0.03, 0.12) | 0.06 (-0.02, 0.14) | 0.04 (-0.04, 0.11) | 0.03 (-0.05, 0.11) |
|  | Common Confounders | 0.12 (-0.19, 0.43) | -0.01 (-0.09, 0.07) | 0.05 (-0.03, 0.13) | 0.05 (-0.02, 0.13) | 0.03 (-0.04, 0.11) | 0.03 (-0.05, 0.11) |
| ASSIST Tobacco | Extended Confounder Set | -0.08 (-0.26, 0.11) | -0.02 (-0.06, 0.03) | -0.01 (-0.06, 0.04) | -0.01 (-0.08, 0.06) | -0.02 (-0.08, 0.03) | -0.02 (-0.06, 0.01) |
|  | Common Confounders | -0.08 (-0.26, 0.09) | -0.02 (-0.06, 0.03) | -0.01 (-0.06, 0.04) | -0.01 (-0.08, 0.06) | -0.02 (-0.08, 0.03) | -0.02 (-0.06, 0.01) |
| ASSIST Alcohol | Extended Confounder Set | -0.08 (-0.26, 0.11) | -0.02 (-0.07, 0.04) | -0.03 (-0.07, 0.02) | -0.01 (-0.08, 0.06) | -0.03 (-0.09, 0.04) | 0.01 (-0.05, 0.06) |
|  | Common Confounders | -0.08 (-0.26, 0.09) | -0.02 (-0.07, 0.04) | -0.03 (-0.07, 0.02) | -0.01 (-0.08, 0.06) | -0.02 (-0.09, 0.04) | 0.01 (-0.05, 0.06) |
| Inverse SES | Extended Confounder Set | 0.63 (-0.02, 1.28) | 0.12 (-0.07, 0.32) | 0.21 (0.02, 0.41) | 0.15 (-0.05, 0.35) | 0.05 (-0.14, 0.24) | 0.19 (-0.01, 0.38) |
|  | Common Confounders | 0.61 (-0.03, 1.24) | 0.11 (-0.08, 0.31) | 0.19 (-0.01, 0.38) | 0.14 (-0.05, 0.34) | 0.05 (-0.13, 0.24) | 0.19 (-0.01, 0.38) |

**Table S5**. Descriptive statistics (Median (IQR)) of prenatal and postnatal indoor air pollutant and psychosocial factor exposures in prenatal and postnatal Self-Organizing Map (SOM) exposure clusters, as seen in Figure 3.

|  | **SOM Cluster** | | | | | |
| --- | --- | --- | --- | --- | --- | --- |
|  | **1** | **2** | **3** | **4** | **5** | **6** |
| N (%) | 129 (21.5) | 32 (5.3) | 86 (14.4) | 134 (22.4) | 69 (11.5) | 149 (24.9) |
| Maternal Age (mean (SD)) | 26.84 (6.02) | 26.98 (5.63) | 26.69 (5.65) | 25.97 (5.65) | 28.14 (5.74) | 27.11 (5.35) |
| Male Child (%) | 59 (45.7) | 16 (50.0) | 45 (52.3) | 82 (61.2) | 38 (55.1) | 69 (46.3) |
| Mixed Ancestry (%) | 15 (11.6) | 9 (28.1) | 64 (74.4) | 85 (63.4) | 23 (33.3) | 77 (51.7) |
| Mother HIV Positive (%) | 42 (32.6) | 10 (31.2) | 14 (16.3) | 21 (15.7) | 25 (36.2) | 22 (14.8) |
| Prenatal Exposures |  |  |  |  |  |  |
| PM10 µg/m3 (median [IQR]) | 41.57 [16.63, 69.29] | 14.86 [3.72, 62.53] | 40.30 [15.29, 72.11] | 41.07 [10.04, 71.14] | 32.10 [16.58, 67.16] | 32.08 [12.07, 67.95] |
| CO mg/m3 (median [IQR]) | 0.00 [0.00, 360.00] | 0.00 [0.00, 0.00] | 0.00 [0.00, 0.00] | 0.00 [0.00, 315.00] | 0.00 [0.00, 500.00] | 0.00 [0.00, 0.00] |
| Benzene µg/m3 (median [IQR]) | 9.16 [3.18, 35.82] | 1.91 [0.44, 7.65] | 3.32 [1.55, 9.62] | 3.78 [1.57, 6.90] | 5.48 [3.11, 40.58] | 3.42 [1.28, 7.00] |
| Toluene µg/m3 (median [IQR]) | 30.19 [14.16, 108.27] | 13.91 [3.47, 41.38] | 16.06 [5.92, 48.55] | 12.59 [5.36, 27.66] | 19.15 [9.47, 57.44] | 14.58 [6.54, 41.59] |
| NO2 µg/m3 (median [IQR]) | 9.25 [4.08, 16.63] | 3.50 [0.00, 6.43] | 7.54 [4.56, 11.38] | 8.05 [3.75, 13.80] | 9.28 [4.34, 15.70] | 6.70 [3.04, 11.00] |
| SO2 µg/m3 (median [IQR]) | 0.00 [0.00, 0.00] | 0.00 [0.00, 0.50] | 0.00 [0.00, 0.18] | 0.00 [0.00, 0.20] | 0.16 [0.00, 0.96] | 0.00 [0.00, 0.28] |
| SES Asset Sum (median [IQR]) | 6.00 [5.00, 7.00] | 7.00 [5.75, 8.00] | 7.00 [5.00, 8.00] | 8.00 [7.00, 8.00] | 7.00 [5.00, 8.00] | 8.00 [7.00, 9.00] |
| Food Insecurity Total Score (median [IQR]) | 3.00 [0.00, 4.00] | 1.00 [0.00, 3.25] | 0.00 [0.00, 2.00] | 0.00 [0.00, 0.00] | 0.00 [0.00, 2.00] | 0.00 [0.00, 0.00] |
| SRQ-20 Total Score (median [IQR]) | 3.00 [1.00, 6.00] | 4.00 [1.75, 6.00] | 6.00 [4.00, 9.00] | 5.00 [3.00, 7.00] | 4.00 [1.00, 6.00] | 2.00 [0.00, 3.00] |
| Emotional IPV Score (median [IQR]) | 4.00 [4.00, 5.00] | 6.00 [4.00, 7.25] | 10.00 [8.00, 13.00] | 5.00 [4.00, 6.00] | 4.00 [4.00, 6.00] | 4.00 [4.00, 5.00] |
| Physical IPV Score (median [IQR]) | 5.00 [5.00, 7.00] | 6.00 [5.00, 10.00] | 11.00 [9.00, 14.00] | 5.00 [5.00, 6.00] | 5.00 [5.00, 6.00] | 5.00 [5.00, 6.00] |
| LEQ Total Score (median [IQR]) | 1.00 [0.00, 1.00] | 1.00 [0.00, 3.00] | 3.00 [2.00, 5.00] | 2.00 [1.00, 4.00] | 2.00 [1.00, 3.00] | 1.00 [0.00, 2.00] |
| EPDS Total Score (median [IQR]) | 11.00 [9.00, 13.00] | 10.00 [6.75, 12.00] | 12.00 [9.00, 16.00] | 10.00 [7.00, 13.00] | 9.00 [7.00, 12.00] | 6.00 [3.00, 9.00] |
| ASSIST Tobacco Score (median [IQR]) | 0.00 [0.00, 0.00] | 0.00 [0.00, 0.00] | 18.50 [0.00, 24.00] | 0.00 [0.00, 23.00] | 0.00 [0.00, 0.00] | 0.00 [0.00, 3.00] |
| ASSIST Alcohol Score (median [IQR]) | 0.00 [0.00, 0.00] | 0.00 [0.00, 0.50] | 0.00 [0.00, 14.75] | 0.00 [0.00, 3.00] | 0.00 [0.00, 0.00] | 0.00 [0.00, 0.00] |
| Postnatal Exposures |  |  |  |  |  |  |
| PM10 µg/m3 (median [IQR]) | 25.35 [13.80, 51.05] | 46.77 [25.77, 78.41] | 25.76 [11.60, 48.31] | 34.89 [16.37, 61.53] | 35.88 [15.03, 75.36] | 24.89 [5.48, 55.72] |
| CO mg/m3 (median [IQR]) | 0.00 [0.00, 60.00] | 60.00 [0.00, 482.50] | 0.00 [0.00, 0.00] | 0.00 [0.00, 0.00] | 0.00 [0.00, 600.00] | 0.00 [0.00, 0.00] |
| Benzene µg/m3 (median [IQR]) | 1.31 [0.49, 4.31] | 1.80 [0.99, 3.36] | 3.47 [0.91, 7.78] | 3.72 [1.60, 6.55] | 95.14 [40.91, 169.71] | 1.89 [0.81, 3.83] |
| Toluene µg/m3 (median [IQR]) | 10.05 [3.15, 18.29] | 10.19 [5.65, 26.44] | 19.74 [9.53, 44.14] | 20.04 [9.92, 47.56] | 405.47 [112.98, 873.22] | 11.33 [5.89, 34.10] |
| NO2 µg/m3 (median [IQR]) | 3.58 [2.05, 7.13] | 6.02 [3.89, 14.60] | 6.45 [3.96, 15.77] | 7.93 [3.52, 14.69] | 10.58 [3.11, 20.46] | 6.09 [3.16, 13.97] |
| SO2 µg/m3 (median [IQR]) | 0.00 [0.00, 0.00] | 9.32 [7.54, 18.29] | 0.00 [0.00, 0.00] | 0.00 [0.00, 0.00] | 0.00 [0.00, 0.00] | 0.00 [0.00, 0.00] |
| SES Asset Sum (median [IQR]) | 7.00 [5.00, 9.00] | 8.00 [6.00, 8.00] | 8.00 [6.00, 9.00] | 8.00 [7.00, 9.00] | 7.00 [5.00, 9.00] | 8.00 [7.00, 9.00] |
| Food Insecurity Total Score (median [IQR]) | 0.00 [0.00, 0.00] | 0.00 [0.00, 0.00] | 0.00 [0.00, 1.00] | 0.00 [0.00, 0.00] | 0.00 [0.00, 0.00] | 0.00 [0.00, 0.00] |
| SRQ-20 Total Score (median [IQR]) | 1.00 [0.00, 3.00] | 0.00 [0.00, 2.25] | 4.00 [2.00, 6.75] | 3.00 [2.00, 6.00] | 3.00 [1.00, 5.00] | 0.00 [0.00, 1.00] |
| Emotional IPV Score (median [IQR]) | 4.00 [4.00, 5.00] | 4.00 [4.00, 5.00] | 9.00 [7.00, 12.00] | 5.00 [4.00, 6.75] | 4.00 [4.00, 6.00] | 4.00 [4.00, 4.00] |
| Physical IPV Score (median [IQR]) | 5.00 [5.00, 6.00] | 5.50 [5.00, 8.00] | 11.00 [7.00, 13.00] | 5.00 [5.00, 7.00] | 5.00 [5.00, 7.00] | 5.00 [5.00, 5.00] |
| LEQ Total Score (median [IQR]) | 0.00 [0.00, 1.00] | 0.50 [0.00, 1.00] | 2.00 [0.25, 4.00] | 2.00 [1.00, 3.00] | 1.00 [0.00, 3.00] | 0.00 [0.00, 1.00] |
| EPDS Total Score (median [IQR]) | 9.00 [7.00, 12.00] | 7.50 [3.50, 10.00] | 10.00 [7.00, 14.00] | 9.00 [5.00, 12.75] | 8.00 [5.00, 13.00] | 4.00 [1.00, 8.00] |
| ASSIST Tobacco Score (median [IQR]) | 0.00 [0.00, 25.00] | 0.00 [0.00, 23.00] | 5.50 [0.00, 25.75] | 6.00 [0.00, 24.00] | 0.00 [0.00, 24.00] | 0.00 [0.00, 2.00] |
| ASSIST Alcohol Score (median [IQR]) | 0.00 [0.00, 0.00] | 0.00 [0.00, 0.75] | 0.00 [0.00, 3.00] | 0.00 [0.00, 6.00] | 0.00 [0.00, 0.00] | 0.00 [0.00, 0.00] |
| Abbreviations: Particulate Matter (PM10); Carbon monoxide (CO); Nitrogen dioxide (NO2); Sulfur dioxide (SO2); Socioeconomic Status (SES); Self-Reporting Questionnaire (SRQ-20); Edinburgh Postnatal Depression Scale (EPDS); Life Experiences Questionnaire (LEQ); Intimate Partner Violence (IPV); Alcohol, Smoking, and Substance Involvement Screening Test (ASSIST) | | | | | | |

**Table S6**. Joint exposure models using combined pre- and postnatal exposure SOM clusters as a joint exposure variables, OR and 95% CIs from adjusted polytomous regression models. Polytomous logistic regression models adjusted for maternal HIV status, maternal age, and ancestry.

|  | Total Problems |
| --- | --- |
|  | Beta (95% CI) |
| Cluster 1 | **0.38 (0.14, 0.62)** |
| Cluster 2 | 0.16 (-0.21, 0.53) |
| Cluster 3 | 0.12 (-0.14, 0.39) |
| Cluster 4 | 0.16 (-0.07, 0.39) |
| Cluster 5 | 0.25 (-0.03, 0.53) |
| Cluster 6 | REF |
|  | Externalizing Problems |
|  | Beta (95% CI) |
| Cluster 1 | **0.29 (0.07, 0.51)** |
| Cluster 2 | 0.01 (-0.34, 0.35) |
| Cluster 3 | 0.17 (-0.07, 0.41) |
| Cluster 4 | 0.16 (-0.05, 0.37) |
| Cluster 5 | 0.24 (-0.02, 0.5) |
| Cluster 6 | REF |
|  | Internalizing Problems |
|  | Beta (95% CI) |
| Cluster 1 | 0.14 (-0.04, 0.31) |
| Cluster 2 | 0 (-0.27, 0.28) |
| Cluster 3 | 0.04 (-0.16, 0.23) |
| Cluster 4 | 0.08 (-0.09, 0.25) |
| Cluster 5 | 0.18 (-0.03, 0.39) |
| Cluster 6 | REF |

**Table S7**. Descriptive statistics (Median (IQR)) of prenatal indoor air pollutant and psychosocial factor exposures in prenatal Self-Organizing Map (SOM) exposure clusters.

|  | **SOM Cluster** | | | |
| --- | --- | --- | --- | --- |
|  | **1** | **2** | **3** | **4** |
| n (%) | 129 (21.5) | 122 (20.4) | 246 (41.1) | 102 (17.0) |
| Maternal Age (mean (SD)) | 28.06 (5.94) | 25.38 (5.12) | 26.76 (5.65) | 27.29 (5.70) |
| Male Child (%) | 64 (49.6) | 72 (59.0) | 122 (49.6) | 51 (50.0) |
| Mixed Ancestry (%) | 32 (24.8) | 108 (88.5) | 65 (26.4) | 68 (66.7) |
| Mother HIV Positive (%) | 48 (37.2) | 13 (10.7) | 55 (22.4) | 18 (17.6) |
| PM10 µg/m3 (median [IQR]) | 38.70 [15.32, 67.16] | 49.06 [13.39, 73.10] | 30.01 [12.09, 62.48] | 34.12 [13.13, 70.24] |
| CO mg/m3 (median [IQR]) | 60.00 [0.00, 850.00] | 0.00 [0.00, 187.50] | 0.00 [0.00, 0.00] | 0.00 [0.00, 0.00] |
| Benzene µg/m3 (median [IQR]) | 36.15 [11.95, 94.02] | 5.25 [2.74, 10.97] | 2.75 [0.90, 4.73] | 3.13 [1.41, 8.24] |
| Toluene µg/m3 (median [IQR]) | 61.16 [27.01, 400.55] | 18.29 [9.42, 54.46] | 10.96 [4.36, 20.50] | 14.21 [5.92, 40.09] |
| NO2 µg/m3 (median [IQR]) | 12.68 [6.69, 19.82] | 9.23 [4.76, 13.17] | 5.40 [2.28, 9.88] | 7.12 [4.34, 11.66] |
| SO2 µg/m3 (median [IQR]) | 0.00 [0.00, 0.43] | 0.00 [0.00, 0.26] | 0.00 [0.00, 0.26] | 0.00 [0.00, 0.23] |
| Urine Cotinine ng/ml (median [IQR]) | 24.80 [10.00, 58.10] | 500.00 [500.00, 500.00] | 17.00 [10.00, 50.88] | 500.00 [37.47, 500.00] |
| SES Asset Sum (median [IQR]) | 6.00 [5.00, 7.00] | 8.00 [7.00, 8.00] | 8.00 [6.00, 8.00] | 7.50 [6.00, 8.00] |
| Food Insecurity Total Score (median [IQR]) | 1.00 [0.00, 4.00] | 0.00 [0.00, 0.00] | 0.00 [0.00, 1.00] | 0.00 [0.00, 2.75] |
| SRQ-20 Total Score (median [IQR]) | 2.00 [1.00, 5.00] | 4.00 [2.00, 7.00] | 3.00 [1.00, 6.00] | 5.50 [4.00, 9.00] |
| Emotional IPV Score (median [IQR]) | 4.00 [4.00, 5.00] | 5.00 [4.00, 6.00] | 4.00 [4.00, 5.00] | 10.00 [8.00, 13.00] |
| Physical IPV Score (median [IQR]) | 5.00 [5.00, 6.00] | 5.00 [5.00, 6.00] | 5.00 [5.00, 6.00] | 11.00 [9.00, 14.00] |
| LEQ Total Score (median [IQR]) | 1.00 [0.00, 2.00] | 2.00 [1.00, 4.00] | 1.00 [0.00, 2.00] | 3.00 [1.00, 5.00] |
| EPDS Total Score (median [IQR]) | 10.00 [7.00, 12.00] | 9.00 [5.00, 12.00] | 9.00 [6.00, 12.00] | 11.00 [8.00, 15.00] |
| ASSIST Tobacco Score (median [IQR]) | 0.00 [0.00, 0.00] | 24.00 [17.25, 25.00] | 0.00 [0.00, 0.00] | 7.50 [0.00, 24.00] |
| ASSIST Alcohol Score (median [IQR]) | 0.00 [0.00, 0.00] | 0.00 [0.00, 6.25] | 0.00 [0.00, 0.00] | 0.00 [0.00, 15.00] |
| Abbreviations: Particulate Matter (PM10); Carbon monoxide (CO); Nitrogen dioxide (NO2); Sulfur dioxide (SO2); Socioeconomic Status (SES); Self-Reporting Questionnaire (SRQ-20); Edinburgh Postnatal Depression Scale (EPDS); Life Experiences Questionnaire (LEQ); Intimate Partner Violence (IPV); Alcohol, Smoking, and Substance Involvement Screening Test (ASSIST) | | | | |

**Table S8.** Joint exposure models using prenatal exposure SOM clusters as a joint exposure variables, OR and 95% CIs from adjusted polytomous regression models. Polytomous logistic regression models adjusted for maternal HIV status, maternal age, and ancestry.

|  | Total Problems |
| --- | --- |
|  | Beta (95% CI) |
| Cluster 1 | 0.18 (-0.03, 0.39) |
| Cluster 2 | **0.34 (0.10, 0.58)** |
| Cluster 3 | REF |
| Cluster 4 | **0.31 (0.07, 0.55)** |
|  | Externalizing Problems |
|  | Beta (95% CI) |
| Cluster 1 | 0.14 (-0.05, 0.33) |
| Cluster 2 | **0.27 (0.05, 0.50)** |
| Cluster 3 | REF |
| Cluster 4 | **0.26 (0.04, 0.48)** |
|  | Internalizing Problems |
|  | Beta (95% CI) |
| Cluster 1 | 0.08 (-0.08, 0.23) |
| Cluster 2 | 0.09 (-0.05, 0.31) |
| Cluster 3 | REF |
| Cluster 4 | 0.09 (-0.06, 0.30) |

**Table S9.** Descriptive statistics (Median (IQR)) of postnatal indoor air pollutant and psychosocial factor exposures in postnatal Self-Organizing Map (SOM) exposure clusters.

|  | **SOM Cluster** | | | |
| --- | --- | --- | --- | --- |
|  | **1** | **2** | **3** | **4** |
| n (%) | 47 (7.8) | 117 (19.5) | 309 (51.6) | 126 (21.0) |
| Maternal Age (mean (SD)) | 27.46 (5.55) | 26.65 (5.81) | 26.53 (5.64) | 27.58 (5.67) |
| Male Child (%) | 22 (46.8) | 63 (53.8) | 150 (48.5) | 74 (58.7) |
| Mixed Ancestry (%) | 13 (27.7) | 74 (63.2) | 122 (39.5) | 64 (50.8) |
| Mother HIV Positive (%) | 11 (23.4) | 25 (21.4) | 66 (21.4) | 32 (25.4) |
| PM10 µg/m3 (median [IQR]) | 41.20 [14.95, 82.34] | 29.28 [13.39, 57.33] | 25.35 [11.12, 51.32] | 35.13 [14.54, 68.44] |
| CO mg/m3 (median [IQR]) | 60.00 [0.00, 485.00] | 0.00 [0.00, 0.00] | 0.00 [0.00, 0.00] | 0.00 [0.00, 505.00] |
| Benzene µg/m3 (median [IQR]) | 1.97 [0.69, 6.34] | 2.69 [0.91, 5.45] | 1.79 [0.72, 3.75] | 32.39 [10.86, 108.78] |
| Toluene µg/m3 (median [IQR]) | 10.95 [6.02, 27.07] | 15.89 [6.52, 35.33] | 11.03 [4.67, 22.65] | 189.77 [55.45, 530.56] |
| NO2 µg/m3 (median [IQR]) | 9.24 [5.05, 14.61] | 6.19 [3.42, 13.90] | 4.75 [2.45, 11.09] | 11.92 [4.75, 18.36] |
| SO2 µg/m3 (median [IQR]) | 7.93 [3.69, 12.62] | 0.00 [0.00, 0.00] | 0.00 [0.00, 0.00] | 0.00 [0.00, 0.00] |
| SES Asset Sum (median [IQR]) | 8.00 [6.00, 9.00] | 8.00 [5.00, 9.00] | 8.00 [6.00, 9.00] | 8.00 [6.00, 9.00] |
| Food Insecurity Total Score (median [IQR]) | 0.00 [0.00, 0.00] | 0.00 [0.00, 1.00] | 0.00 [0.00, 0.00] | 0.00 [0.00, 0.00] |
| SRQ-20 Total Score (median [IQR]) | 0.00 [0.00, 2.00] | 5.00 [2.00, 9.00] | 1.00 [0.00, 2.00] | 3.00 [1.00, 5.00] |
| Emotional IPV Score (median [IQR]) | 4.00 [4.00, 4.00] | 9.00 [7.00, 12.00] | 4.00 [4.00, 4.00] | 4.00 [4.00, 6.00] |
| Physical IPV Score (median [IQR]) | 5.00 [5.00, 7.00] | 11.00 [7.00, 13.00] | 5.00 [5.00, 5.00] | 5.00 [5.00, 6.00] |
| LEQ Total Score (median [IQR]) | 0.00 [0.00, 1.00] | 2.00 [1.00, 4.00] | 0.00 [0.00, 2.00] | 1.00 [0.00, 3.00] |
| EPDS Total Score (median [IQR]) | 8.00 [5.00, 10.00] | 11.00 [7.00, 16.00] | 7.00 [4.00, 10.00] | 8.00 [5.00, 13.00] |
| ASSIST Tobacco Score (median [IQR]) | 0.00 [0.00, 21.00] | 16.00 [0.00, 26.00] | 0.00 [0.00, 16.00] | 3.00 [0.00, 23.25] |
| ASSIST Alcohol Score (median [IQR]) | 0.00 [0.00, 0.00] | 0.00 [0.00, 3.00] | 0.00 [0.00, 0.00] | 0.00 [0.00, 3.00] |
| Abbreviations: Particulate Matter (PM10); Carbon monoxide (CO); Nitrogen dioxide (NO2); Sulfur dioxide (SO2); Socioeconomic Status (SES); Self-Reporting Questionnaire (SRQ-20); Edinburgh Postnatal Depression Scale (EPDS); Life Experiences Questionnaire (LEQ); Intimate Partner Violence (IPV); Alcohol, Smoking, and Substance Involvement Screening Test (ASSIST) | | | | |

**Table S10.** Joint exposure models using postnatal exposure SOM clusters as a joint exposure variables, OR and 95% CIs from adjusted polytomous regression models. Polytomous logistic regression models adjusted for maternal HIV status, maternal age, and ancestry.

|  | Total Problems |
| --- | --- |
|  | Beta (95% CI) |
| Cluster 1 | -0.16 (-0.46, 0.14) |
| Cluster 2 | -0.08 (-0.29, 0.13) |
| Cluster 3 | REF |
| Cluster 4 | 0.05 (0.26, 0.15) |
|  | Externalizing Problems |
|  | Beta (95% CI) |
| Cluster 1 | -0.14 (-0.42, 0.13) |
| Cluster 2 | 0.01 (-0.19, 0.20) |
| Cluster 3 | REF |
| Cluster 4 | 0.06 (-0.13, 0.24) |
|  | Internalizing Problems |
|  | Beta (95% CI) |
| Cluster 1 | -0.22 (-0.44, 0.01) |
| Cluster 2 | -0.08 (-0.24, 0.08) |
| Cluster 3 | REF |
| Cluster 4 | -0.04 (-0.19, 0.11) |

**Figures**

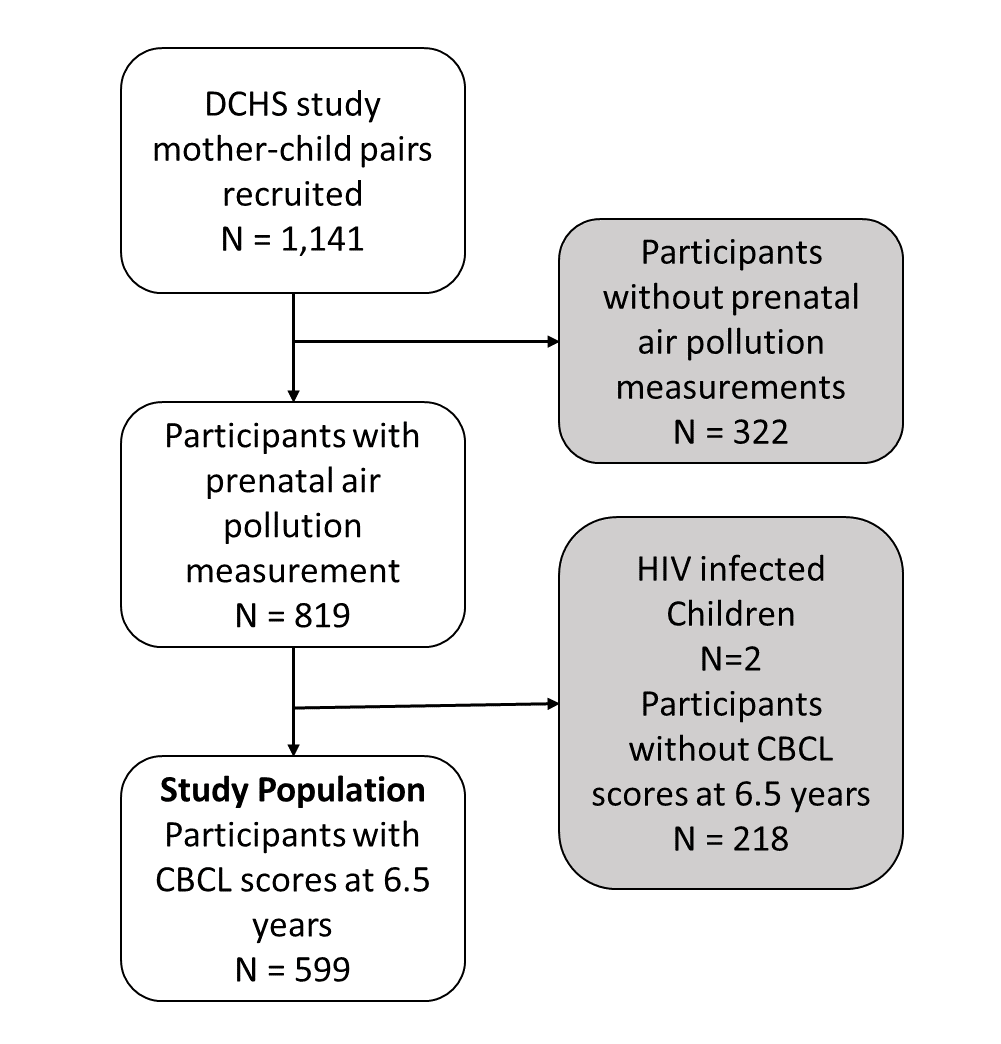

Figure S1. Study population flow diagram.

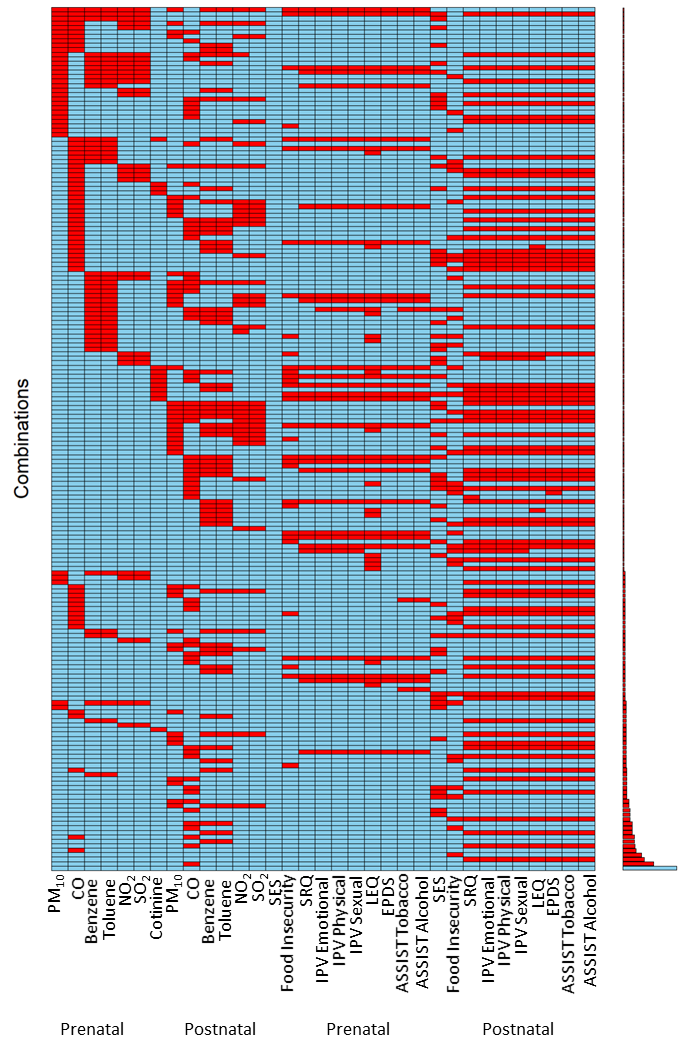

**Figure S2.** Combinations of missingness patterns of exposure variables. Each row is a missingness pattern where red indicates that variable is missing, and blue indicates not missing. Abbreviations: Particulate Matter (PM10); Carbon monoxide (CO); Nitrogen dioxide (NO2); Sulfur dioxide (SO2); Socioeconomic Status (SES); Self-Reporting Questionnaire (SRQ-20); Edinburgh Postnatal Depression Scale (EPDS); Life Experiences Questionnaire (LEQ); Intimate Partner Violence (IPV); Alcohol, Smoking, and Substance Involvement Screening Test (ASSIST).

A.

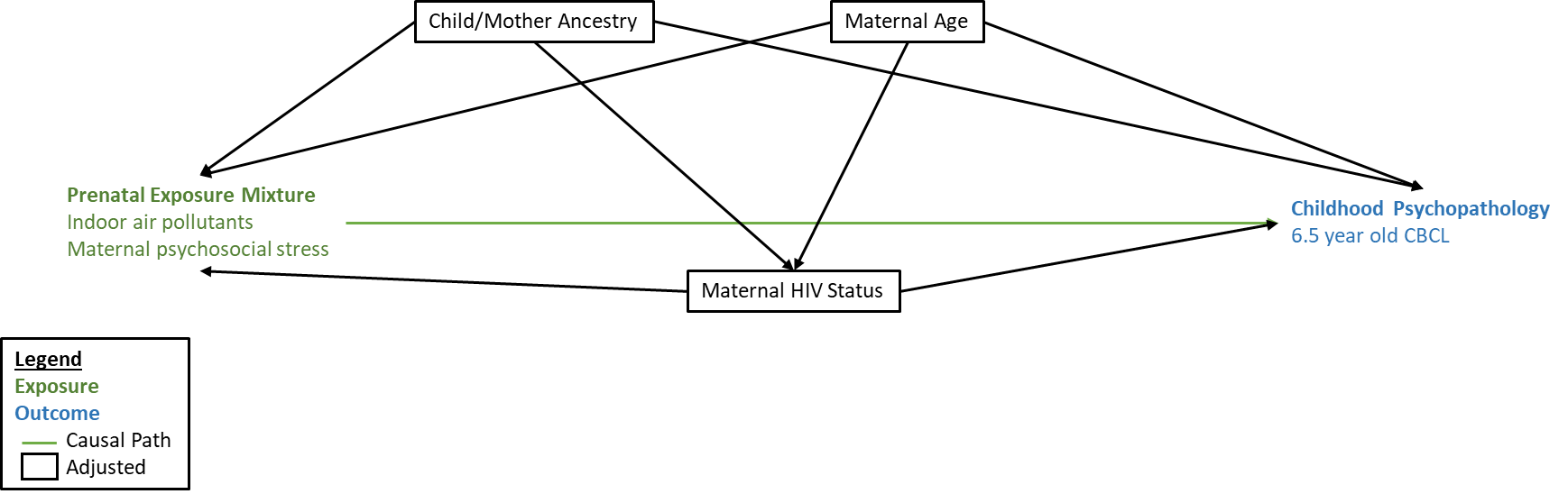

B.

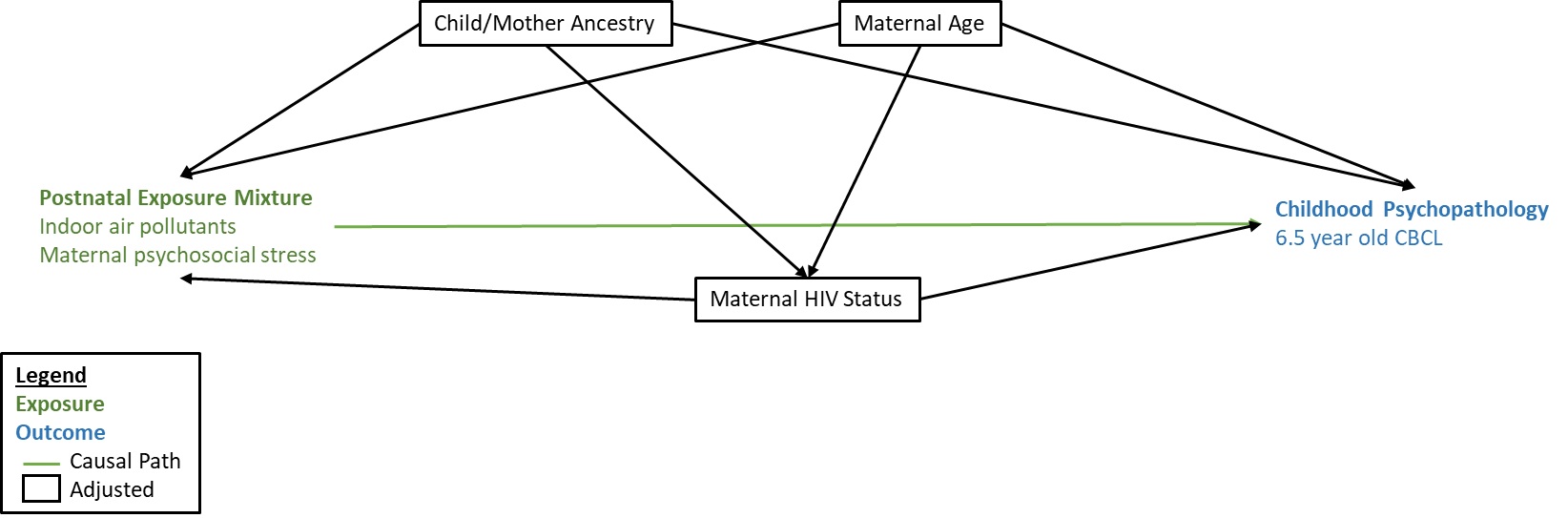

C.

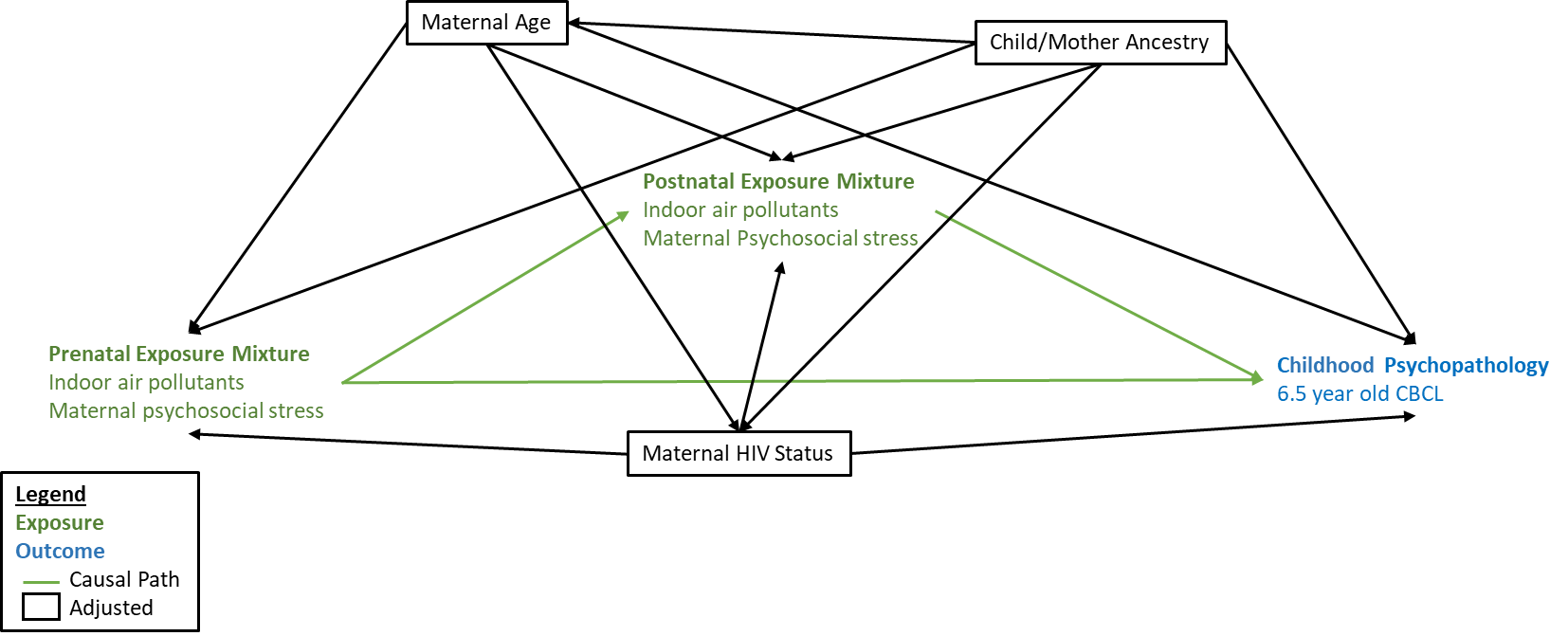

**Figure S3. DAGs of underlying causal pathways between pre- and postnatal exposure to indoor air pollutants and psychosocial factors including socioeconomic status, and childhood psychopathology at 6.5 years**. A. DAG for prenatal exposure only. B. DAG for postnatal exposure only. C. DAG for pre- and postnatal exposure.

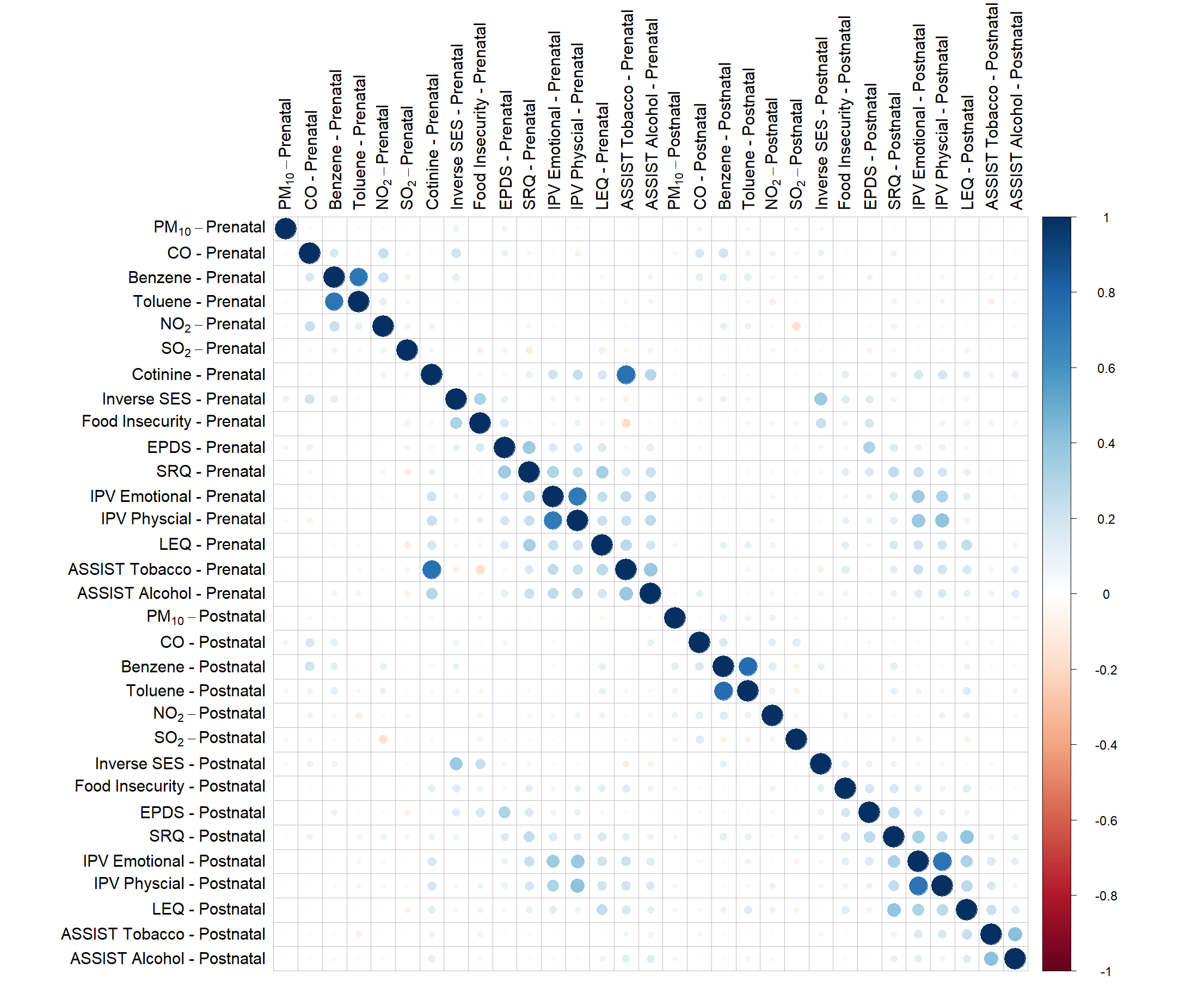

**Figure S4**. Pearson correlation matrix with pre- and postnatal exposure measurements. Abbreviations: Particulate Matter (PM10); Carbon monoxide (CO); Nitrogen dioxide (NO2); Sulfur dioxide (SO2); Socioeconomic Status (SES); Self-Reporting Questionnaire (SRQ-20); Edinburgh Postnatal Depression Scale (EPDS); Life Experiences Questionnaire (LEQ); Intimate Partner Violence (IPV); Alcohol, Smoking, and Substance Involvement Screening Test (ASSIST).

**A. Prenatal exposure with CBCL total problems. B. Prenatal exposure with CBCL externalizing problems. C. Prenatal exposure with CBCL internalizing problems**

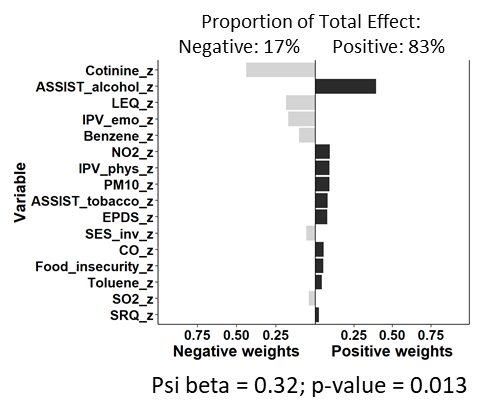

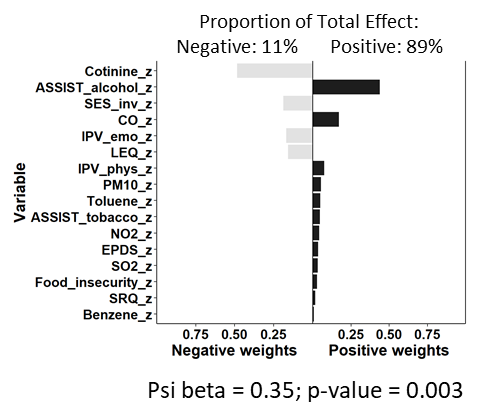

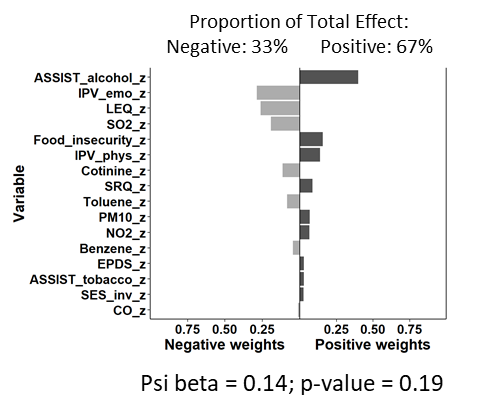

**D. Postnatal exposure with CBCL total problems. E. Postnatal exposure with CBCL externalizing problems. F. Postnatal exposure with CBCL internalizing problems**

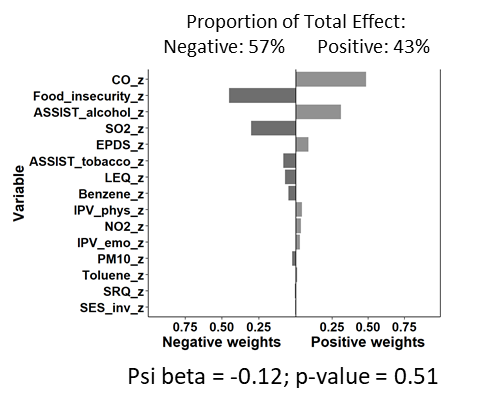

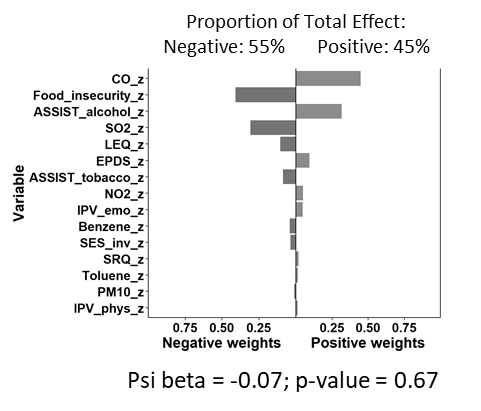

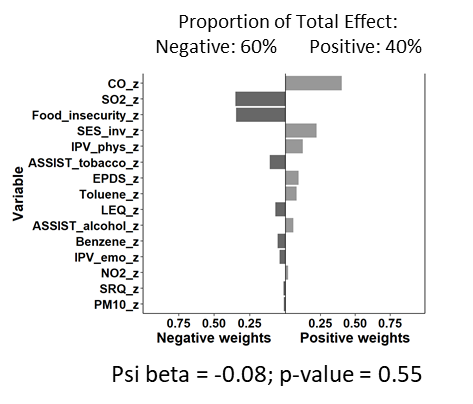

**Figure S5. Weight of each exposure mixture component from quantile G-computation models, adjusted for maternal age, maternal HIV status, and ancestry. Weights from models were the total mixture effect was not significant should not be interpreted**. A. Association between prenatal exposure mixture and CBCL total problems. B. Association between prenatal exposure mixture and CBCL externalizing problems. C. Association between prenatal exposure mixture and CBCL internalizing problems. D. Association between postnatal exposure mixture and CBCL total problems. E. Association between postnatal exposure mixture and CBCL externalizing problems. F. Association between postnatal exposure mixture and CBCL internalizing problems. Abbreviations: Particulate Matter (PM10); Carbon monoxide (CO); Nitrogen dioxide (NO2); Sulfur dioxide (SO2); Socioeconomic Status (SES); Self-Reporting Questionnaire (SRQ-20); Edinburgh Postnatal Depression Scale (EPDS); Life Experiences Questionnaire (LEQ); Intimate Partner Violence (IPV); Alcohol, Smoking, and Substance Involvement Screening Test (ASSIST).

A.	B.

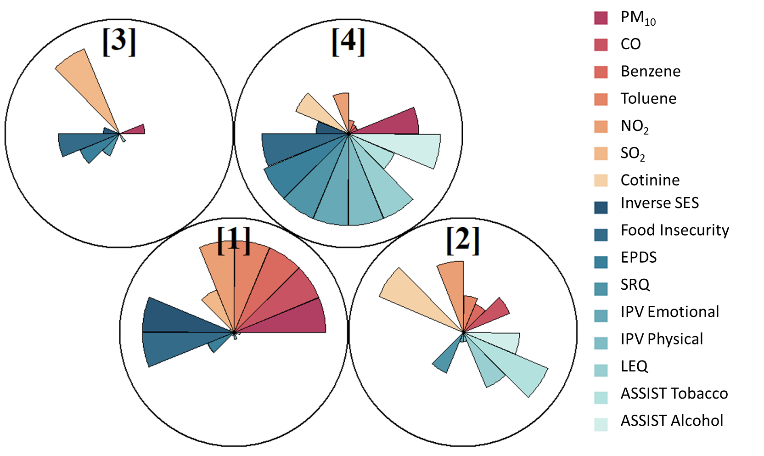

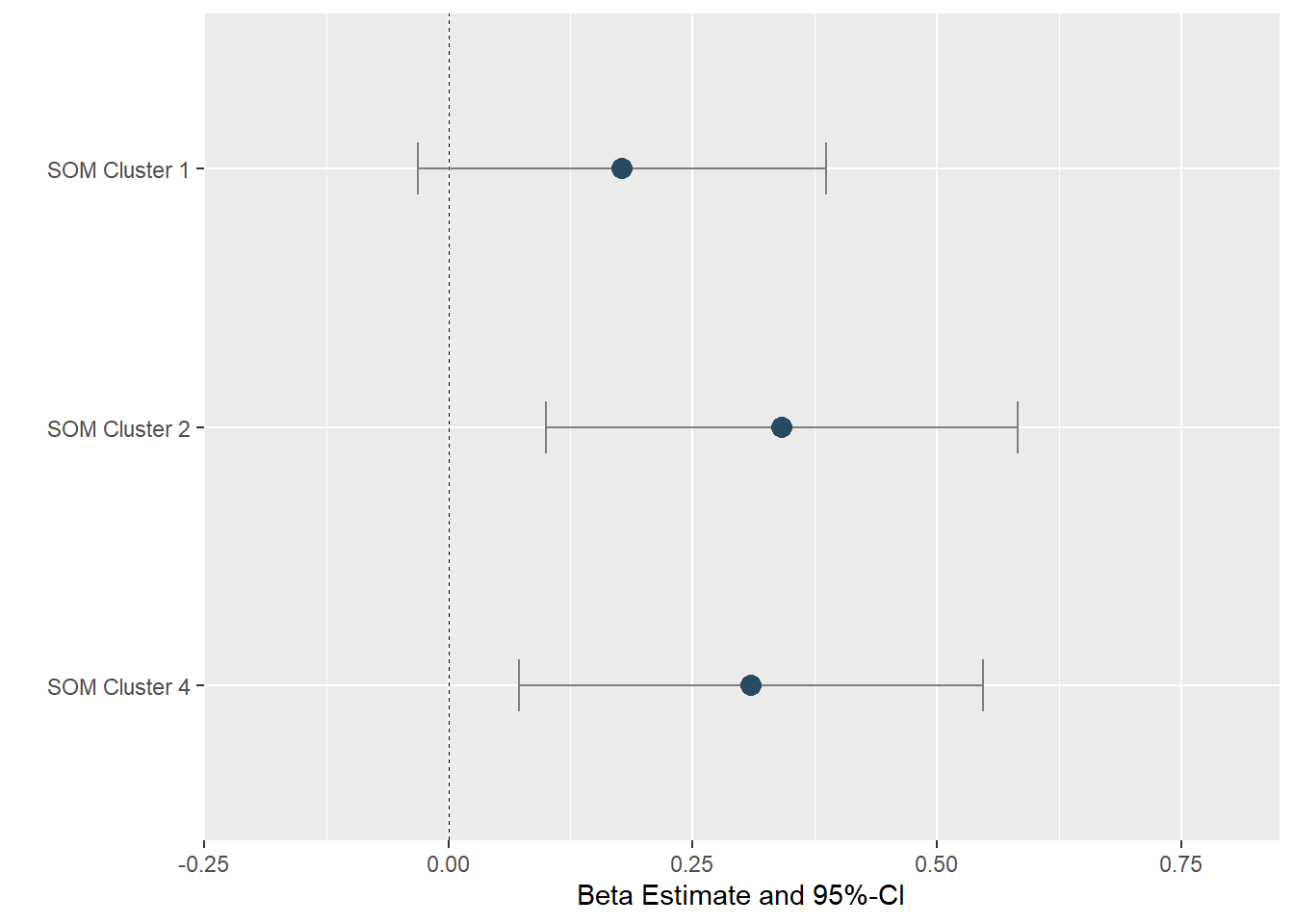

C.	D.

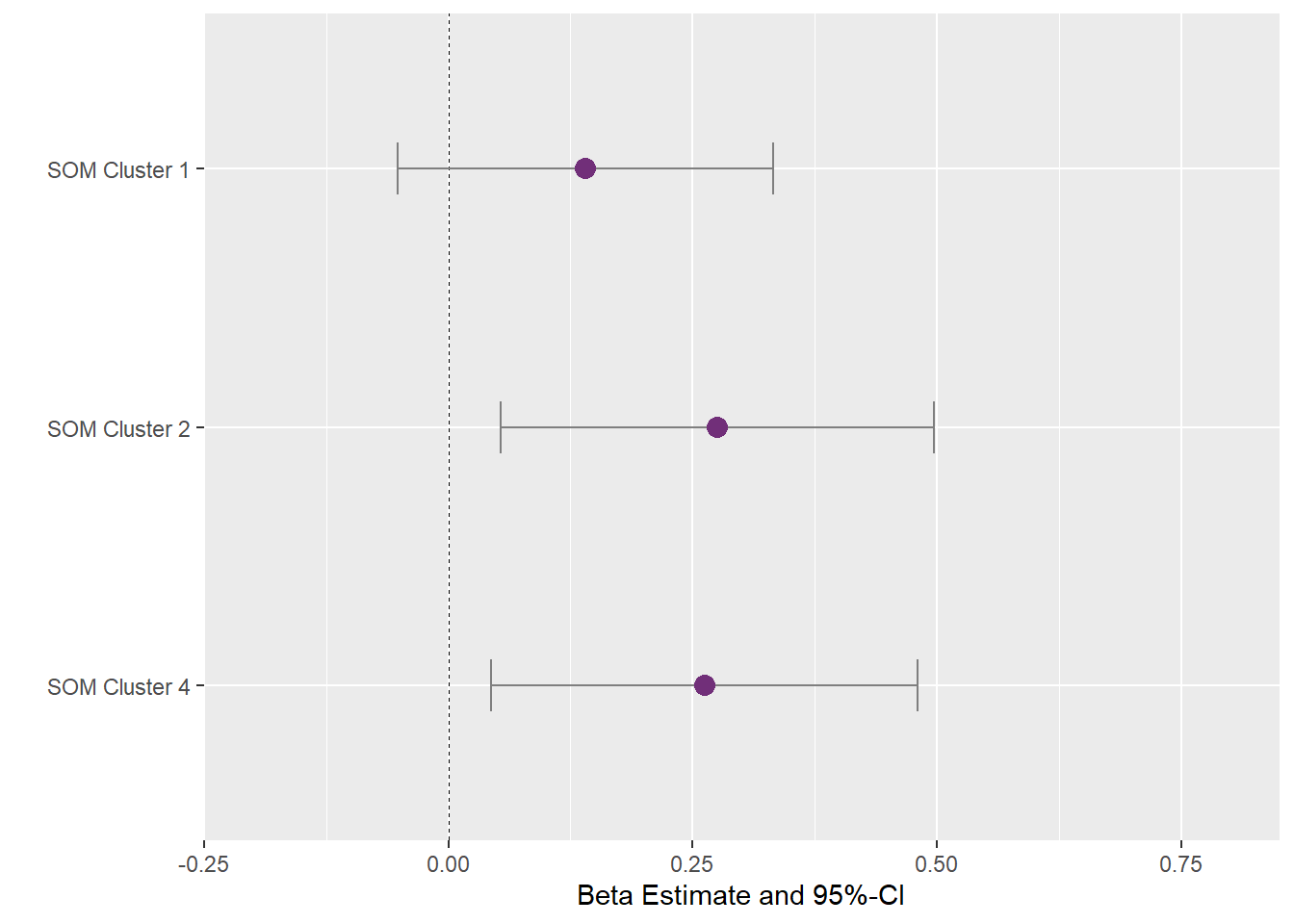

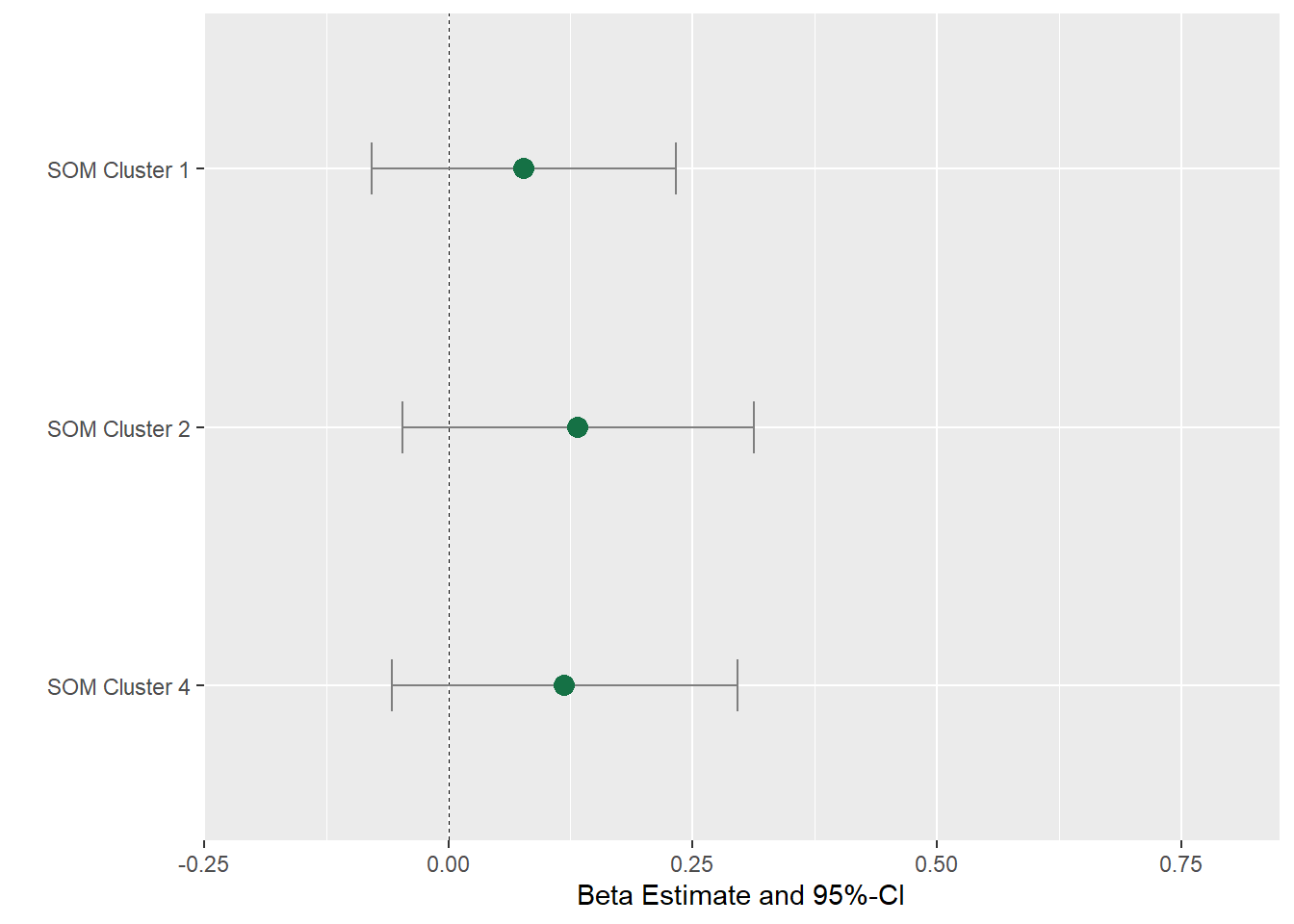

**Figure S6. Results from self-organizing map (SOM) analysis using prenatal indoor air pollutants and psychosocial factors.** A. SOM clusters created using pre- natal indoor air pollutants and psychosocial factors. B. Associations between SOM clusters and CBCL total problems, adjusted by maternal age, maternal HIV status, and ancestry. SOM cluster 3 is used as the reference group. C. Associations between SOM clusters and CBCL externalizing problems, adjusted by maternal age, maternal HIV status, and ancestry. SOM cluster 3 is used as the reference group. D. Associations between SOM clusters and CBCL internalizing problems, adjusted by maternal age, maternal HIV status, and ancestry. SOM cluster 3 is used as the reference group. Abbreviations: Particulate Matter (PM10); Carbon monoxide (CO); Nitrogen dioxide (NO2); Sulfur dioxide (SO2); Socioeconomic Status (SES); Self-Reporting Questionnaire (SRQ-20); Edinburgh Postnatal Depression Scale (EPDS); Life Experiences Questionnaire (LEQ); Intimate Partner Violence (IPV); Alcohol, Smoking, and Substance Involvement Screening Test (ASSIST).

A.	B.

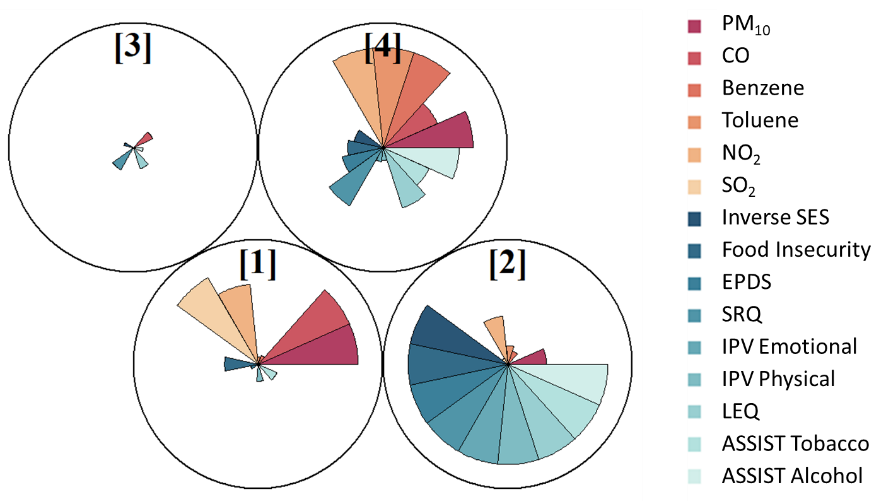

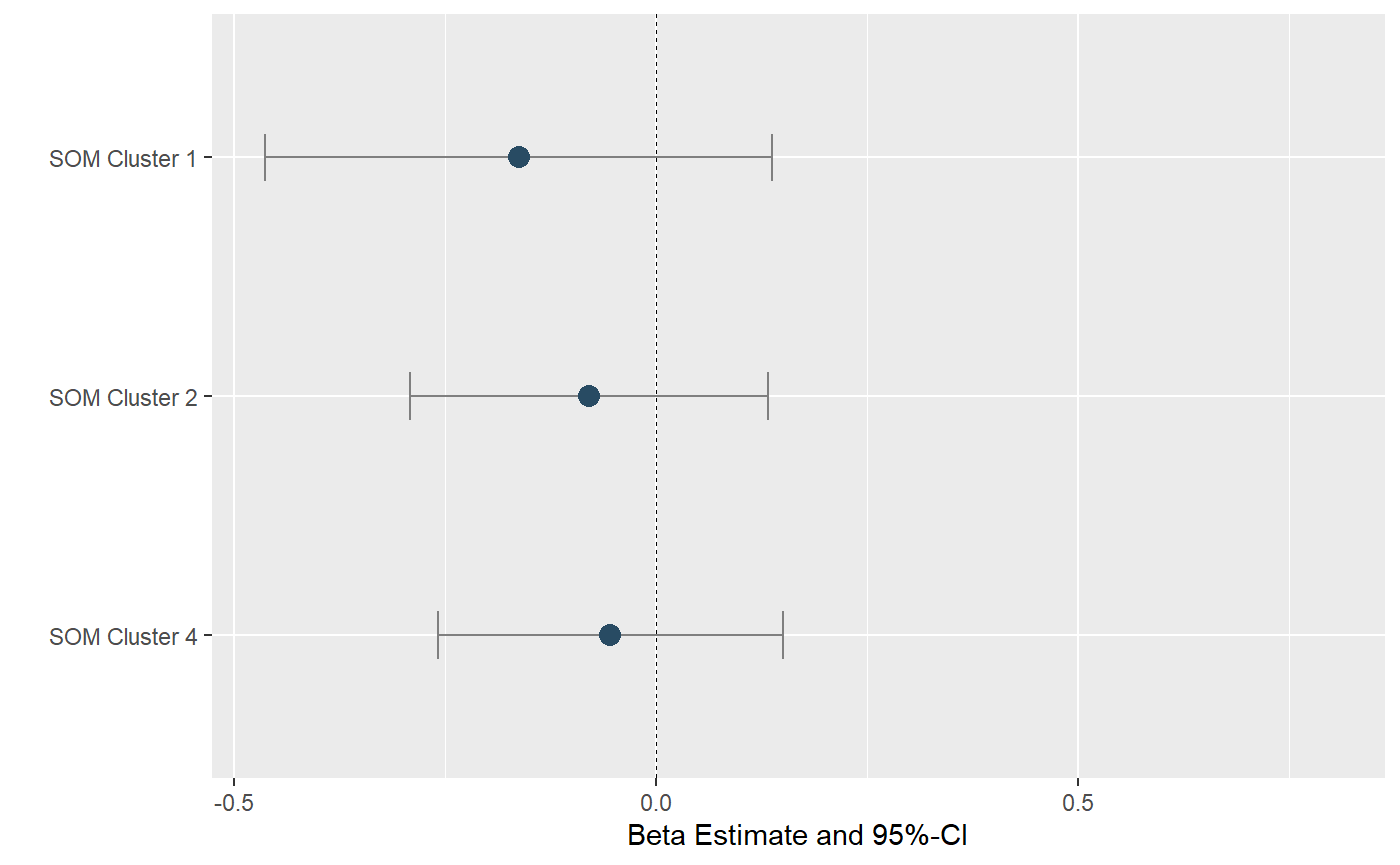

C.	D.

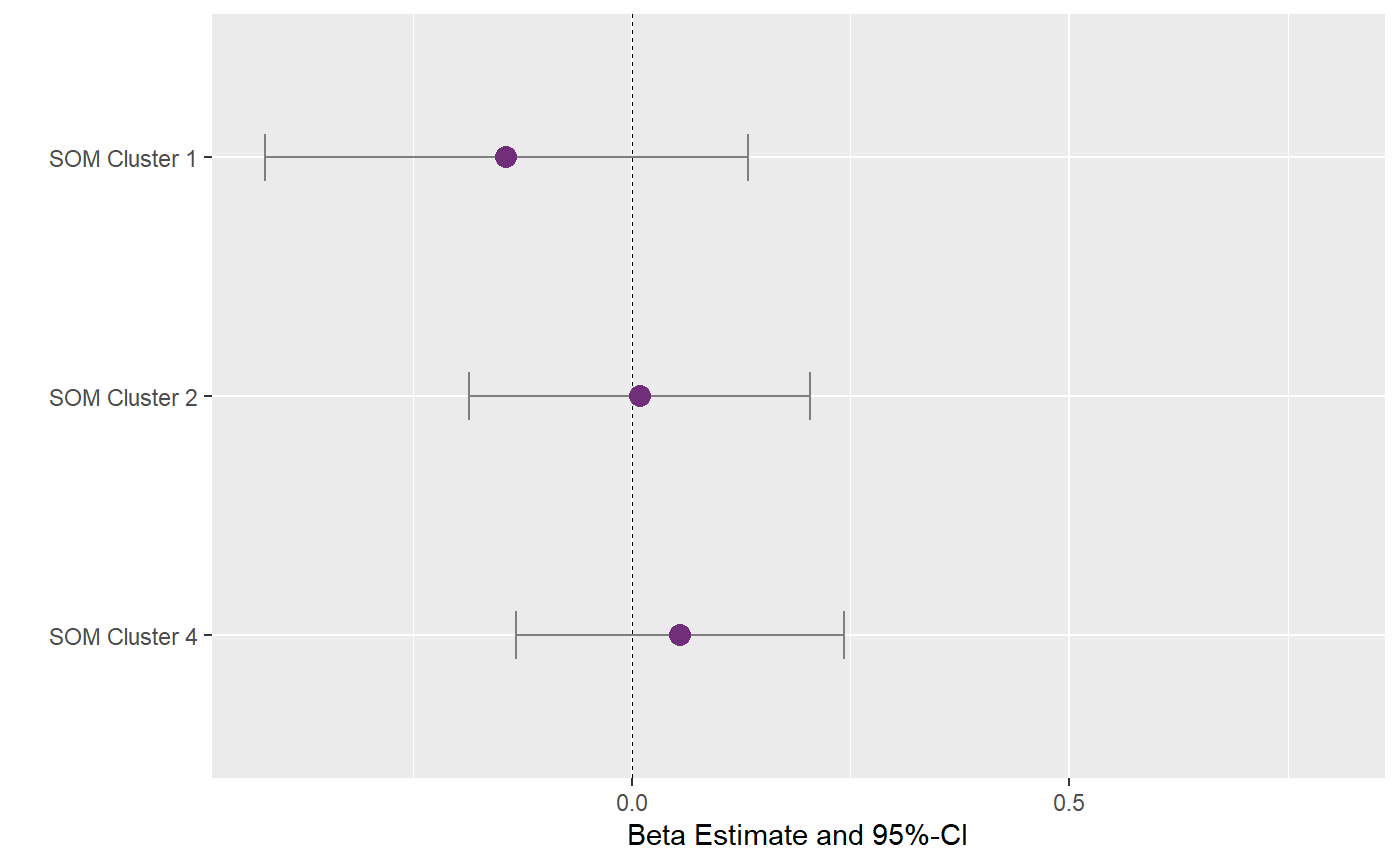

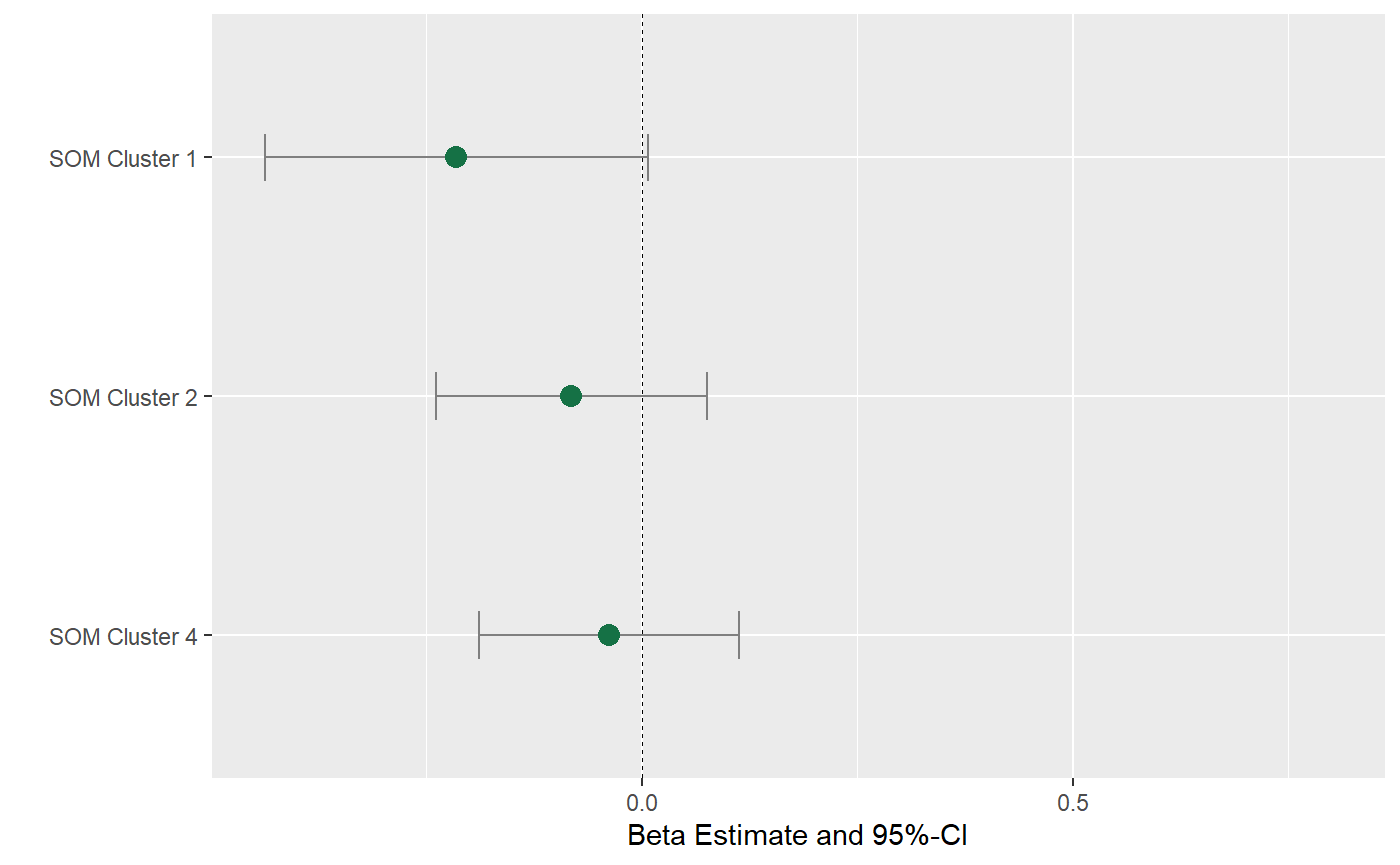

**Figure S7. Results from self-organizing map (SOM) analysis using postnatal indoor air pollutants and psychosocial factors.** A. SOM clusters created using postnatal indoor air pollutants and psychosocial factors. B. Associations between SOM clusters and CBCL total problems, adjusted by maternal age, maternal HIV status, and ancestry. SOM cluster 3 is used as the reference group. C. Associations between SOM clusters and CBCL externalizing problems, adjusted by maternal age, maternal HIV status, and ancestry. SOM cluster 3 is used as the reference group. D. Associations between SOM clusters and CBCL internalizing problems, adjusted by maternal age, maternal HIV status, and ancestry. SOM cluster 3 is used as the reference group. Abbreviations: Particulate Matter (PM10); Carbon monoxide (CO); Nitrogen dioxide (NO2); Sulfur dioxide (SO2); Socioeconomic Status (SES); Self-Reporting Questionnaire (SRQ-20); Edinburgh Postnatal Depression Scale (EPDS); Life Experiences Questionnaire (LEQ); Intimate Partner Violence (IPV); Alcohol, Smoking, and Substance Involvement Screening Test (ASSIST).
